# Sperm from infertile, oligozoospermic men have elevated mutation rates

**DOI:** 10.1101/2024.08.22.24312232

**Authors:** Jason Kunisaki, Michael E. Goldberg, Suchita Lulla, Thomas Sasani, Laurel Hiatt, Thomas J. Nicholas, Lihua Liu, Elizabeth Torres-Arce, Yixuan Guo, Emma James, Joshua J Horns, Joemy M Ramsay, Qi Chen, James M Hotaling, Kenneth I Aston, Aaron R. Quinlan

## Abstract

Male infertility is associated with elevated rates of aneuploidy and DNA breaks in spermatozoa and germline precursors. This common condition is not well understood and is associated with poor individual and familial somatic health relative to fertile men. To further understand the extent and source of genome instability, we used error-corrected duplex DNA sequencing to test whether the impaired spermatogenesis and relatively poorer health of oligozoospermic men are linked to elevated single nucleotide *de novo* mutation frequencies in their sperm and blood, respectively. We observed a significant 1.34 to 2.01-fold increase in age-adjusted sperm mutation frequencies in infertile, oligozoospermic men. Conversely, consistently elevated mutation frequencies in the blood of oligozoospermic men were not found. Gain-of-function mutations linked to clonal spermatogenesis and Mendelian disorders accumulate with age at a similar rate in normozoospermic and oligozoospermic men. These results implicate germline hypermutation as a hallmark feature of oligozoospermia and point to age-independent processes affecting spermatogonial stem cell biology that may underlie spermatogenic impairment before and after puberty. Our findings also underscore the importance of investigating tissue-specific mechanisms driving the association between reduced reproductive and somatic health in infertile men.

## Introduction

Human male fertility is a heterogeneous trait with broad diversity in quantitative and qualitative phenotypes^1–3^. For example, normozoospermic (i.e., normal sperm production) men have sperm concentrations ranging from >15 million to hundreds of millions of sperm per milliliter of ejaculate^4^. In contrast, azoospermic and oligozoospermic men present with sperm concentrations ranging from 0 to less than 15 million sperm per milliliter, respectively. Beyond differences in sperm concentration, many fertile men with qualitatively normal semen parameters, including motility, morphology, and viability, may have up to 70% sperm with impaired motility and 96% with abnormal morphology^5,6^.

This phenotypic variability mirrors the complexity of sperm production, which is mediated by exogenous and endogenous factors that affect the abundance and function of spermatogonial stem cells (SSCs). Smoking^7^, chemotherapy^8^, and pesticide exposure^9^ each reduce SSC viability and impair their ability to produce sperm. Genetic factors, including Y chromosome microdeletions^10^ and sex chromosome aneuploidies^11^, contribute to spermatogenic failure and explain ∼30% of male infertility cases^12^. Genome-wide association studies of non-obstructive azoospermic (NOA, zero sperm in ejaculate) and severe oligozoospermic (SO, <5 million sperm/mL) cases have identified only a handful of genes potentially implicated in reduced SSC function and impaired spermatogenesis^13–18^. Recent exome sequencing studies using large cohorts of NOA and SO men have had greater success, identifying dozens of causal variants, genes, and pathways underlying monogenic forms of spermatogenic dysfunction^19–23^. However, these associations are often derived from few individuals and limited to severe, largely recessive phenotypes. Consequently, while these studies uncover the genetic heterogeneity underlying reduced SSC function and spermatogenic impairment, their findings have yet to be broadly integrated into clinical management strategies for infertile men^24^, as 75% of NOA and 90% of oligozoospermic cases remain idiopathic^25^. As such, the full range of molecular etiologies underlying male infertility—a condition that affects ∼7% of men worldwide—has yet to be discovered.

Despite the genetic heterogeneity of male infertility, large-scale forms of genetic instability, including aneuploidies^26–29^, chromosomal microdeletions^30^, and fragmented DNA^31^, are consistently observed in the testes and sperm of infertile men. These clinical findings suggest that maintaining genomic integrity is critical for SSC self-renewal and differentiation^32,33^. For example, a large body of evidence from mouse models suggests that the inactivation of DNA repair genes involved in mismatch repair, chromosomal synapsis, and meiotic recombination contribute to increased aneuploidy rates and spermatogenic failure^34–38^. Nevertheless, the factors connecting SSC dysfunction and subsequent germline mutagenesis in humans remain elusive, in part because human SSCs cannot be cultured *in vitro*^*39*^.

In addition to reproductive complications, infertile men often present with unexplained poor somatic health. Numerous epidemiologic studies report varying effect sizes for the association of male infertility with shorter lifespan^40,41^, and several studies have demonstrated the relationship between male infertility and increased risks for genital cancers^42–46^ and cardiovascular disease^47,48^. In a prior study of germline mutations in large, multigenerational pedigrees, we found that men in the highest quartile of age-adjusted germline *de novo* mutation (DNM) rates live, on average, nearly five fewer years than those in the lowest quartile^49^. Following the somatic theory of aging, which posits that mutations accumulate over time and contribute to morbidity and mortality^50^, this finding suggests that higher germline mutation rates may be a biomarker for elevated somatic mutation burden and poorer health.

Genome sequencing of parent-child trios has been the standard approach for investigating rates and patterns of germline DNMs in parental gametes and, by extension, helped characterize germline development^51–58^. However, relying on pedigrees to study germline development and mutagenesis typically excludes infertile men incapable of reproduction via natural pregnancies. Sequencing bulk sperm, which even in fertile men is composed of reproductively fit and unfit gametes^59^, may help overcome these biases and provide a deeper understanding of male germline development and genetic heterogeneity across male fertility phenotypes. Recent studies, for example, have applied duplex sequencing^60–63^ and single-sperm genome sequencing^64^ to directly examine germline mutations from bulk sperm.

This study builds upon these efforts by investigating how spermatogenic impairment and poor health in oligozoospermia are associated with DNM rates and patterns. Specifically, we use error-corrected, duplex DNA sequencing to analyze low-frequency, single-nucleotide *de novo* mutations (DNMs) in sperm and blood from oligozoospermic men, allowing us to explore potential associations between genomic instability, impaired spermatogenesis, and poor somatic health in infertile men. We test whether spermatogenic impairment in oligozoospermic men is associated with elevated sperm mutation rates. To explore a potential explanation for the epidemiologic associations between male infertility and poor health, we also investigate whether increased germline mutation rates predict elevated blood mutation rates in oligozoospermic men.

## Results

### Mutation detection in bulk sperm and blood with duplex DNA sequencing

Detecting rare mutations present in sperm or blood cells is inherently difficult since the error rates of modern DNA sequencing technologies are orders of magnitude higher than mutation rates. For example, the average male germline mutation rate is estimated to be ∼0.83×10^-8^ per base pair per gamete^52,53,55^, while errors in modern DNA sequencing technologies (e.g., Illumina) arise, on average, roughly once for every 1,000 nucleotides sequenced^65^. Duplex sequencing addresses this signal-to-noise problem by distinguishing true mutations as those present on complementary DNA strands from sequencing and PCR errors that arise on single strands. A “duplex consensus” read (“duplex read,” in short) is inferred from the amplified reads from each strand and attenuates errors to a theoretical rate of between 1 in 10 million^66^ and 1 in a billion^60^. Because each duplex consensus read represents a single DNA fragment derived from a haploid gamete, mutations detected on a consensus-generated read can be ascribed to a single haploid sperm cell.

By sequencing samples to an intended duplex molecule depth of up to 8,000X, on average, we can detect mutations at a given nucleotide site among 8,000 distinct sperm cells. However, because short DNA fragments are barcoded from a population of gametes, multiple mutations present in a single sperm cell cannot be tracked to the same gamete unless they are each observed in the same duplex molecule. It is cost-prohibitive to sequence the entire human genome at such high depth with duplex sequencing; therefore, we designed a 325 kilobase targeted sequencing panel covering coding sequences of 93 genes relevant for sperm production, genomic integrity, and somatic health (“custom” panel). These genes were selected based on their involvement in aspects of male fertility, including DNA repair, clonal spermatogenesis, cancer, and monogenic drivers of infertility (**Methods, Supplementary Table 1**). Furthermore, we targeted a 48kb panel consisting of 20 control loci (“control panel”), used in prior studies to estimate genome-wide mutation frequencies and spectra in mice^67,68^ and humans^69^.

Deep duplex DNA sequencing was obtained across the custom and control regions (∼373 kilobases in total) to identify mutations found in at least one cell from bulk sperm and matched blood samples among normozoospermic and oligozoospermic donors. Donor samples were provided by the Subfertility Health Assisted Reproduction and the Environment cohort, a large biobank at the University of Utah, with matched sperm and blood samples from more than 7,000 donors. All normozoospermic donors had clinically normal sperm concentrations above 15 million/mL of ejaculate and comprised a “cross-sectional” and a “longitudinal” cohort. The longitudinal cohort includes ten normozoospermic men with proven fertility who had sperm samples collected at two distinct time points with 12 to 24 years between collection dates (average of 19.1 years). Blood was collected from longitudinal donors at the second time point. While two donors in this cohort exhibited low sperm concentrations at their 2nd time point (NL5 - 9.7 M/mL and NL10 - 9.0 M/mL, **Figure 1, Table 1**), semen analyses from these donors 2 to 3 weeks later reported sperm concentrations within ranges of normozoospermia **(Supplementary Table 2**). The cross-sectional cohort includes 15 normozoospermic donors, each of whom was part of a male-female couple seeking fertility counseling from the University of Utah Andrology Clinic. Semen samples from the cross-sectional cohort were clinically normal, including sperm concentration, suggesting male factor infertility was not a major contributor in each infertility case (**Methods**). Conversely, oligozoospermic men possessed abnormally low sperm concentrations upon standard diagnostic workups at the Andrology Clinic, which likely contributed to their infertility (**Figure 1, Table 1**).

**Figure 1.**
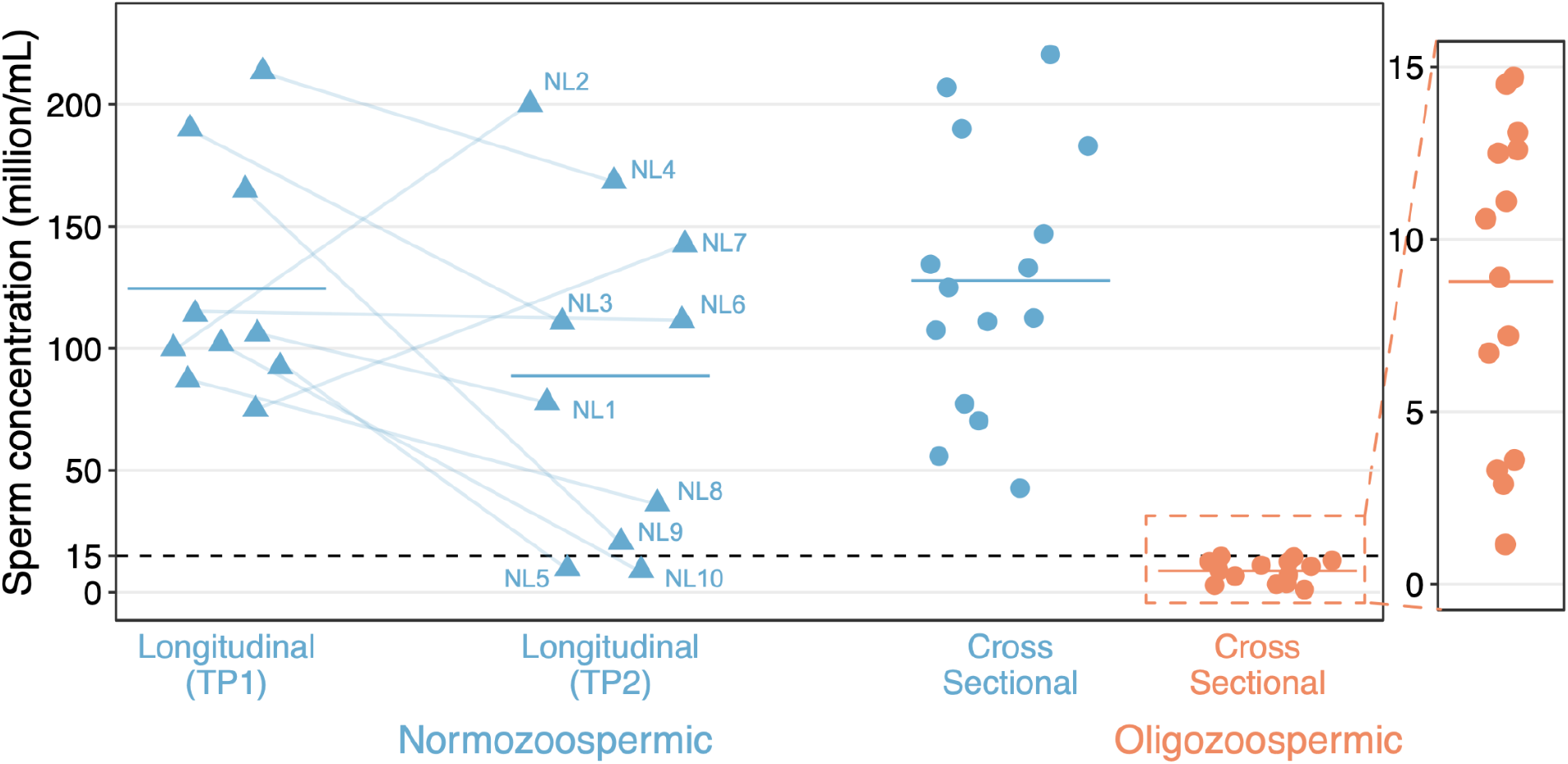
Duplex sequencing to study DNMs in bulk sperm from a unique cohort of normozoospermic and oligozoospermic men. Sperm concentrations (million sperm per mL of semen) for normozoospermic (blue) and oligozoospermic (orange) men are depicted in each cohort. Longitudinally collected sperm samples from normozoospermic men are marked as triangles. For each donor, a blue line connects data points corresponding to sperm concentrations measured at the 1st time point (TP1) and the 2nd time point (TP2). The solid lines within each group indicate median sperm concentrations. The dashed horizontal black line highlights the sperm concentration cutoff of 15M sperm/mL. The enlarged orange plot highlights the range of sperm concentrations (all <15M sperm/mL) specifically for the oligozoospermic cohort.

**Table 1.**
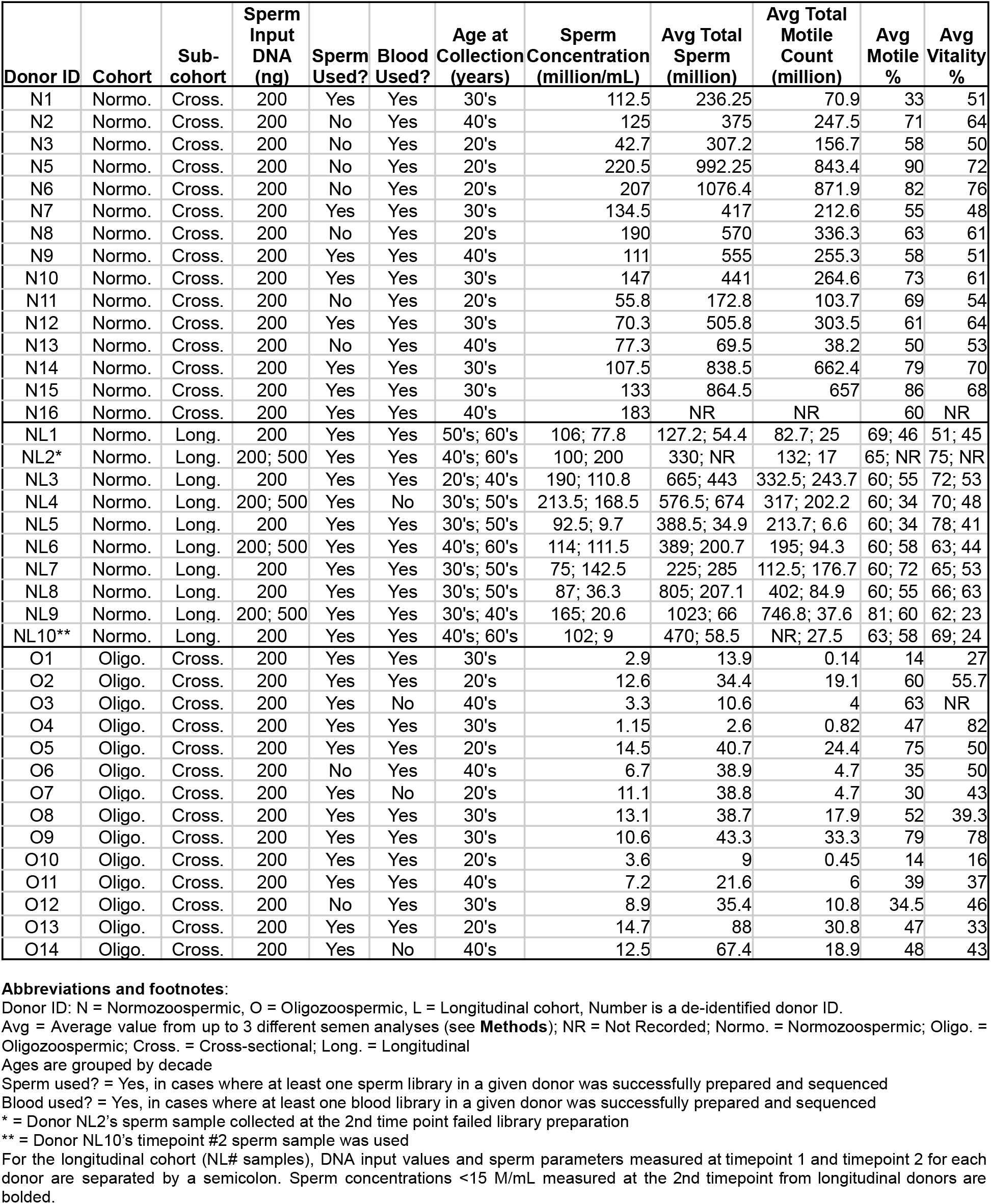
Summary of normozoospermic and oligozoospermic sperm donors.

| Donor ID | Cohort | Sub-cohort | Sperm Input DNA (ng) | Sperm Used? | Blood Used? | Age at Collection (years) | Sperm Concentration (million/mL) | Avg Total Sperm (million) | Avg Total Motile Count (million) | Avg Motile % | Avg Vitality % |
| --- | --- | --- | --- | --- | --- | --- | --- | --- | --- | --- | --- |
| N1 | Normo. | Cross. | 200 | Yes | Yes | 30's | 112.5 | 236.25 | 70.9 | 33 | 51 |
| N2 | Normo. | Cross. | 200 | No | Yes | 40's | 125 | 375 | 247.5 | 71 | 64 |
| N3 | Normo. | Cross. | 200 | No | Yes | 20's | 42.7 | 307.2 | 156.7 | 58 | 50 |
| N5 | Normo. | Cross. | 200 | No | Yes | 20's | 220.5 | 992.25 | 843.4 | 90 | 72 |
| N6 | Normo. | Cross. | 200 | No | Yes | 20's | 207 | 1076.4 | 871.9 | 82 | 76 |
| N7 | Normo. | Cross. | 200 | Yes | Yes | 30's | 134.5 | 417 | 212.6 | 55 | 48 |
| N8 | Normo. | Cross. | 200 | No | Yes | 20's | 190 | 570 | 336.3 | 63 | 61 |
| N9 | Normo. | Cross. | 200 | Yes | Yes | 40's | 111 | 555 | 255.3 | 58 | 51 |
| N10 | Normo. | Cross. | 200 | Yes | Yes | 30's | 147 | 441 | 264.6 | 73 | 61 |
| N11 | Normo. | Cross. | 200 | No | Yes | 20's | 55.8 | 172.8 | 103.7 | 69 | 54 |
| N12 | Normo. | Cross. | 200 | Yes | Yes | 30's | 70.3 | 505.8 | 303.5 | 61 | 64 |
| N13 | Normo. | Cross. | 200 | No | Yes | 40's | 77.3 | 69.5 | 38.2 | 50 | 53 |
| N14 | Normo. | Cross. | 200 | Yes | Yes | 30's | 107.5 | 838.5 | 662.4 | 79 | 70 |
| N15 | Normo. | Cross. | 200 | Yes | Yes | 30's | 133 | 864.5 | 657 | 86 | 68 |
| N16 | Normo. | Cross. | 200 | Yes | Yes | 40's | 183 | NR | NR | 60 | NR |
| NL1 | Normo. | Long. | 200 | Yes | Yes | 50's; 60's | 106; 77.8 | 127.2; 54.4 | 82.7; 25 | 69; 46 | 51; 45 |
| NL2* | Normo. | Long. | 200; 500 | Yes | Yes | 40's; 60's | 100; 200 | 330; NR | 132; 17 | 65; NR | 75; NR |
| NL3 | Normo. | Long. | 200 | Yes | Yes | 20's; 40's | 190; 110.8 | 665; 443 | 332.5; 243.7 | 60; 55 | 72; 53 |
| NL4 | Normo. | Long. | 200; 500 | Yes | No | 30's; 50's | 213.5; 168.5 | 576.5; 674 | 317; 202.2 | 60; 34 | 70; 48 |
| NL5 | Normo. | Long. | 200 | Yes | Yes | 30's; 50's | 92.5; 9.7 | 388.5; 34.9 | 213.7; 6.6 | 60; 34 | 78; 41 |
| NL6 | Normo. | Long. | 200; 500 | Yes | Yes | 40's; 60's | 114; 111.5 | 389; 200.7 | 195; 94.3 | 60; 58 | 63; 44 |
| NL7 | Normo. | Long. | 200 | Yes | Yes | 30's; 50's | 75; 142.5 | 225; 285 | 112.5; 176.7 | 60; 72 | 65; 53 |
| NL8 | Normo. | Long. | 200 | Yes | Yes | 30's; 50's | 87; 36.3 | 805; 207.1 | 402; 84.9 | 60; 55 | 66; 63 |
| NL9 | Normo. | Long. | 200; 500 | Yes | Yes | 30's; 40's | 165; 20.6 | 1023; 66 | 746.8; 37.6 | 81; 60 | 62; 23 |
| NL10** | Normo. | Long. | 200 | Yes | Yes | 40's; 60's | 102; 9 | 470; 58.5 | NR; 27.5 | 63; 58 | 69; 24 |
| O1 | Oligo. | Cross. | 200 | Yes | Yes | 30's | 2.9 | 13.9 | 0.14 | 14 | 27 |
| O2 | Oligo. | Cross. | 200 | Yes | Yes | 20's | 12.6 | 34.4 | 19.1 | 60 | 55.7 |
| O3 | Oligo. | Cross. | 200 | Yes | No | 40's | 3.3 | 10.6 | 4 | 63 | NR |
| O4 | Oligo. | Cross. | 200 | Yes | Yes | 30's | 1.15 | 2.6 | 0.82 | 47 | 82 |
| O5 | Oligo. | Cross. | 200 | Yes | Yes | 20's | 14.5 | 40.7 | 24.4 | 75 | 50 |
| O6 | Oligo. | Cross. | 200 | No | Yes | 40's | 6.7 | 38.9 | 4.7 | 35 | 50 |
| O7 | Oligo. | Cross. | 200 | Yes | No | 20's | 11.1 | 38.8 | 4.7 | 30 | 43 |
| O8 | Oligo. | Cross. | 200 | Yes | Yes | 30's | 13.1 | 38.7 | 17.9 | 52 | 39.3 |
| O9 | Oligo. | Cross. | 200 | Yes | Yes | 30's | 10.6 | 43.3 | 33.3 | 79 | 78 |
| O10 | Oligo. | Cross. | 200 | Yes | Yes | 20's | 3.6 | 9 | 0.45 | 14 | 16 |
| O11 | Oligo. | Cross. | 200 | Yes | Yes | 40's | 7.2 | 21.6 | 6 | 39 | 37 |
| O12 | Oligo. | Cross. | 200 | No | Yes | 30's | 8.9 | 35.4 | 10.8 | 34.5 | 46 |
| O13 | Oligo. | Cross. | 200 | Yes | Yes | 20's | 14.7 | 88 | 30.8 | 47 | 33 |
| O14 | Oligo. | Cross. | 200 | Yes | No | 40's | 12.5 | 67.4 | 18.9 | 48 | 43 |

**Abbreviations and footnotes:**
Donor ID: N = Normozoospermic, O = Oligozoospermic, L = Longitudinal cohort, Number is a de-identified donor ID.
Avg = Average value from up to 3 different semen analyses (see **Methods**); NR = Not Recorded; Normo. = Normozoospermic; Oligo. = Oligozoospermic; Cross. = Cross-sectional; Long. = Longitudinal
Ages are grouped by decade
Sperm used? = Yes, in cases where at least one sperm library in a given donor was successfully prepared and sequenced
Blood used? = Yes, in cases where at least one blood library in a given donor was successfully prepared and sequenced
\* = Donor NL2's sperm sample collected at the 2nd time point failed library preparation
\*\* = Donor NL10's timepoint #2 sperm sample was used
For the longitudinal cohort (NL# samples), DNA input values and sperm parameters measured at timepoint 1 and timepoint 2 for each donor are separated by a semicolon. Sperm concentrations <15 M/mL measured at the 2nd timepoint from longitudinal donors are bolded.

We imposed strict bioinformatic filters on the duplex DNA sequencing data from bulk sperm and blood samples to detect high-confidence mutations. We removed low-quality duplex consensus reads, bases, and mutations called at sites in error-prone regions of the panel (**Methods**), excluding an average of ∼23.6% (SD = 0.5%) of duplex bases and ∼81.9% (SD = 7.1%) of mutation calls in normozoospermic and oligozoospermic donors. These filters resulted in a median duplex depth of ∼6,500x and ∼4,000x from 200ng DNA libraries derived from the tissues of normozoospermic and oligozoospermic men, respectively (**Supplementary Figure 1**). The proportion of low-quality bases and mutations that were excluded was similar across tissues and samples from both cohorts, indicating that error rates were consistent across samples (**Supplementary Tables 3 and 4**). Sequencing thousands of sperm at specific genomic regions theoretically allowed us to distinguish nonclonal mutations found in a single duplex read from clonal mutations inferred to be present in multiple gametes based on their detection in two or more duplex reads. However, even severely oligozoospermic men can possess millions of sperm in their ejaculate. Therefore, it is likely that many rare yet clonal mutations exist at frequencies below the limits of our detection and are only detected in a single gamete or never observed^61,70^. Most (>94%) detected mutations post-filter were found on a single duplex read and, therefore, attributed to a single cell (**Supplementary Figure 2** and **Supplementary Table 5**).

We calculated each sample’s mutation frequency by dividing its count of unique, high-confidence mutations by the total number of high-quality bases sequenced (**Supplementary Tables 5 and 6**). By examining technical replicates of sperm and blood samples from normozoospermic and oligozoospermic men, we found that each sample’s mutation frequency is consistently measured using our duplex sequencing approach (**Methods, Supplementary Figure 3**). Downsampling experiments further demonstrated that observed mutation frequencies are robust to differences in total sequencing depth between normozoospermic and oligozoospermic donors (**Supplementary Figure 4**). Donors whose sperm or blood library preparation failed (**Methods**) may prevent the exclusion of mutations found in both tissue types for a given donor. Nevertheless, we demonstrate that our rigorous set of filters can identify DNMs that would be flagged as recurrent in matched tissues, yielding consistent mutation frequency estimates with or without a successfully prepared matched tissue sample (**Methods, Supplementary Figure 5**).

### Mutation frequencies increase with age in the same donors

Numerous pedigree studies of germline mutations have observed a strong paternal age effect. While an average of 1.5 additional mutations are observed per sperm each year a man ages, we recently found that this effect varies among fathers, with mutations accumulating at rates between 0.19 to nearly 3.24 additional mutations per year^52^. As a positive control, we tested for a paternal age effect in the eight longitudinal donors who had library preparations of their bulk sperm at two distinct time points. As expected, we observed a significant increase in mutation frequency with age (Poisson regression with donor interaction term *P =* 5.82×10^-6^); on average, the male germlines in our cohort accumulated an average of 2.87 DNMs/year (**Figure 2**). While this increase with age is roughly 1.91-fold greater than the average rate of mutation accumulation observed in pedigree studies, the variability in germline mutation accumulation rates across the longitudinal cohort reflects the family-specific variability we previously observed from genome sequencing 603 individuals from 33 large multi-generation families^52^. Indeed, an ANOVA goodness-of-fit test reports a significantly better fit when including donor ID and its interaction with age in the Poisson model (ANOVA *P* = 5.078×10^-79^), highlighting how donor-specific factors could influence germline mutation accumulation rates with increased age. Furthermore, the rate of mutation accumulation with age was consistent among 200ng and 500ng DNA replicates for three donors with successful sperm libraries (NL4, NL6, and NL9), suggesting accurate measures of paternal age effect in the longitudinal cohort. We observed a negative age effect in donors NL3 and NL9, likely due to the sampling of sperm lineages that acquired fewer mutations by the second collection time point relative to lineages sampled at the first time point.

**Figure 2.**
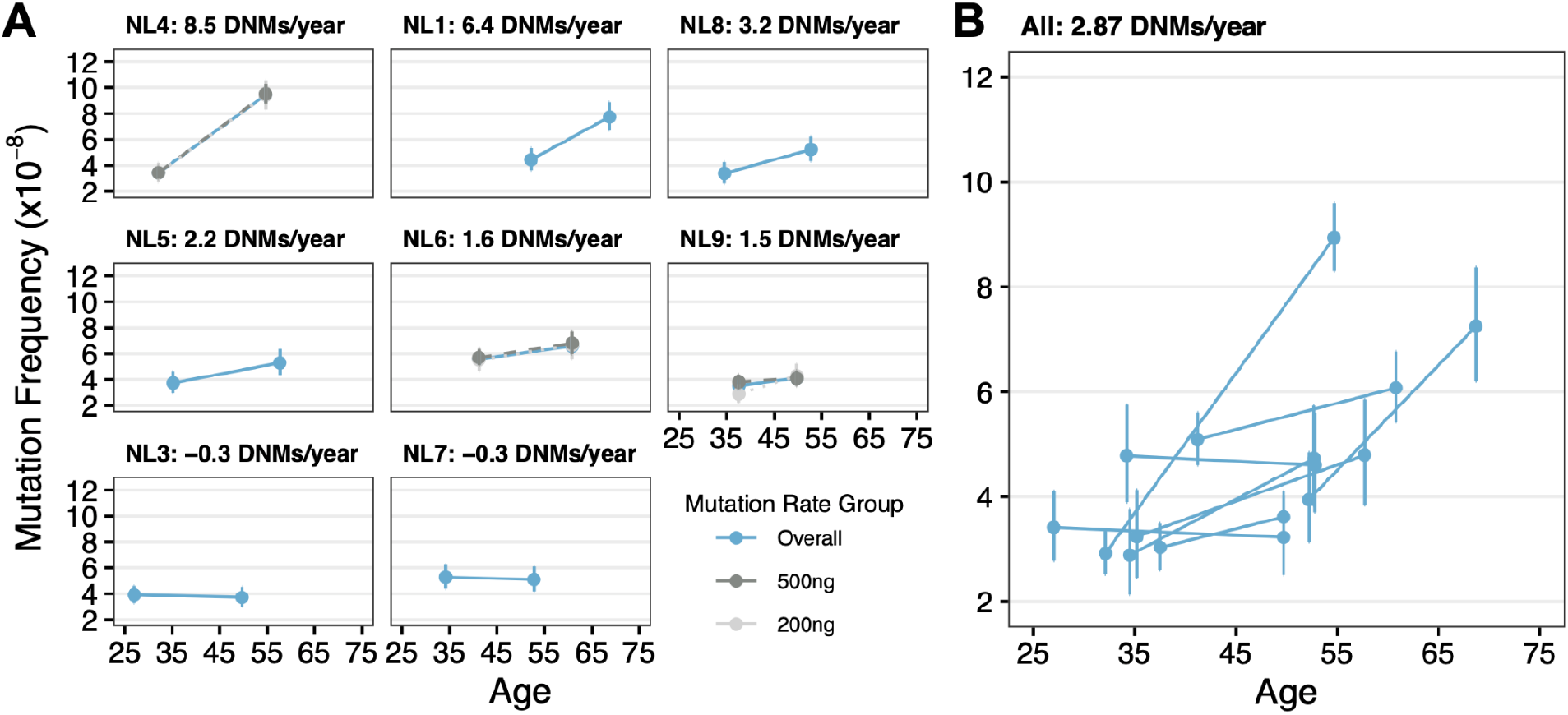
Germline mutations increase with age in longitudinally sampled bulk sperm from normozoospermic donors. **A)** Mutation frequencies in bulk sperm samples collected at two time points from 8 longitudinal donors. Each panel depicts mutation frequencies and 95% confidence intervals measured at the donor’s age at 1st collection (left) and 2nd collection (right). The donors are sorted in descending order by their rate of mutation accumulation per year. Donors with technical bulk sperm replicates prepared with 200ng (light gray) and 500ng (darker gray) of DNA are plotted separately. Blue data points reflect overall mutation frequencies measured from bulk sperm at each time point. **B)** The overall mutation frequencies measured at two time points from each donor are plotted as individual lines. The overall rate of mutation accumulation was calculated as the average rate from all eight donors.

### Oligozoospermic men exhibit elevated sperm mutation frequencies relative to normozoospermic men

Based on prior observations of increased aneuploidy and DNA breakage in the sperm of oligozoospermic men, we hypothesized that their sperm would also exhibit elevated single-nucleotide mutation frequencies under the assumption that DNA damage accumulates in their spermatogonial stem cells. To test this hypothesis, we first established baseline estimates for sperm mutation frequencies in normozoospermic men. For longitudinal donors with multiple semen samples available, we analyzed sperm collected at the second time point to restrict our analysis to independent samples. Furthermore, given the consistency in mutation frequencies estimated from technical replicates derived from 200ng and 500ng DNA libraries, we restricted our analysis to 200ng samples, the input used across all oligozoospermic samples. Because our data was overdispersed when modeled with a Poisson regression (**Methods**), we instead modeled mutation counts in normozoospermic and oligozoospermic men using a negative binomial regression. These models employed a log link function and included an offset term to directly account for variability in duplex coverage across samples (**Methods**). With an average age of 47 years old in the normozoospermic cohort, our model reported mutation frequencies of 4.58×10^-8^/duplex bp (**Figure 3A**, 95% CI: 4.03×10^-8^/duplex bp to 5.14×10^-8^/duplex bp). As expected, our negative binomial regression model identified a significant increase in mutation frequency with age (*P* = 4.06×10^-2^). Bae et al.^63^ recently applied duplex sequencing to bulk sperm from a 39-year-old male and reported a sperm mutation frequency of 2.7×10^-8^/duplex bp. After adjusting for paternal age, their finding is roughly 1.47-fold lower than our model’s estimate; however, Bae et al. analyzed bulk sperm from a single donor. Comparing our results to a study that used duplex sequencing to report sperm mutation frequencies of 2.5×10^-8^ in a slightly larger cohort of six 18-year-old males^60^, our predicted mutation frequencies of 2.76×10^-8^/duplex bp (95% CI: 1.86×10^-8^/duplex bp to 3.67×10^-8^/duplex bp) at this age are in line with previous work.

**Figure 3.**
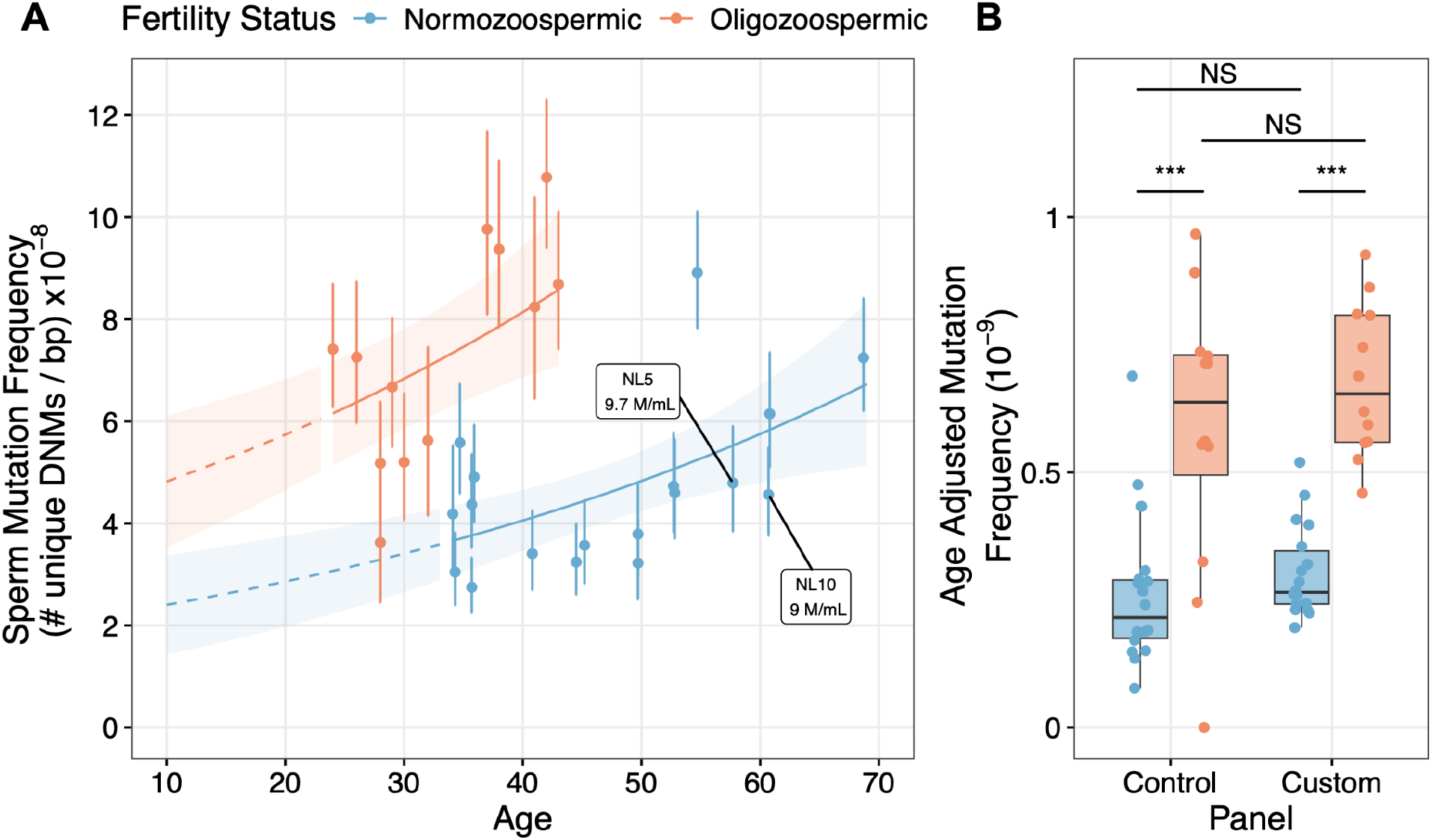
Oligozoospermic men exhibit elevated mutation frequencies in their sperm. **A)** Mutation frequencies measured in normozoospermic (blue) and oligozoospermic (orange) men. The shaded portion indicates the standard error around the mutation frequency estimates. The regression model generated from the observed data points was used to predict the mutation frequencies at unobserved ages between 10 (start of puberty) and 70 (dotted line). **B)** Age-adjusted mutation frequencies (mutation frequency / donor age) are shown when subsetting to control and custom panel mutations. NS indicates a permutation test p-value > 0.05. An *** indicates a significant permutation test p-value < 0.01 (**Methods**).

Our estimates of germline mutation frequencies build upon prior studies that sequenced the genomes of parent-child trios to investigate germline mutations in parental gametes^58^ and demonstrate that sperm progenitor cells accumulate DNMs with age and throughout the embryonic development of the father^52,55,57^. However, family studies strictly examine germline mutations in the subset of sperm capable of fertilization. Therefore, our current understanding of germline mutation is heavily biased towards reproductively fit sperm from fertile men. Our measures of mutations in bulk sperm collected from normozoospermic and oligozoospermic men address the biases in pedigree studies by directly detecting mutations from reproductively fit and unfit gametes in a bulk cell population. This difference in biological source material (i.e., bulk sperm vs reproductively fit sperm) may underlie increased mutation frequency estimates measured from bulk sperm relative to values reported by pedigree studies. Alternatively, errors in duplex sequencing may artificially inflate mutation frequency estimates from bulk sperm relative to pedigree-based approaches that use whole-genome sequencing. Using a strict set of filters to identify a high-confidence set of mutations, we found that error rates are consistent across samples (**Supplementary Tables 2 and 3**), suggesting that any observed differences in mutation frequencies in the sperm of normozoospermic and oligozoospermic men can be attributed to a biological rather than technical source.

Given that error rates were consistent across samples and cohorts, we compared the mutation frequencies observed in sperm samples from normozoospermic and oligozoospermic men. We observed a higher average mutation frequency of 7.20×10^-8^/duplex bp (**Figure 3A**, 95% CI: 6.19×10^-8^/duplex bp to 8.22×10^-8^/duplex bp) in the oligozoospermic cohort, despite having an average age of 33 years old, which is 14 years younger than the average age in the normozoospermic cohort. We also found that oligozoospermic men ≤35 years old have statistically similar mutation frequencies to estimates from normozoospermic men older than 50 years old (Welch two sample t-test *P* = 0.5015 vs *P* = 0.01389 when compared to normozoospermic men ≤35 years old).

To further characterize the relationship between germline mutagenesis and male subfertility, we included age and fertility status as covariates in our model of mutation frequencies. The average sperm motility in each donor’s semen sample was also included as a covariate to correct for the possibility that immotile sperm, which have been shown to exhibit increased aneuploidy rates compared to motile sperm^71^, may also harbor elevated single nucleotide DNM burdens. With these considerations, our negative binomial model identified a significant increase in sperm mutation frequencies measured from oligozoospermic men (**Methods**, negative binomial *P* = 2.97×10^-8^), corresponding to a 2.01-fold increase in mutation frequency at the start of puberty. We found that including an interaction term between age and fertility status to model potential differences in age-dependent mutation accumulation in oligozoospermic men did not improve the model’s fit (ANOVA *P* = 0.2301), suggesting that mutation accumulation rates with age were similar across our cohorts. We also observed a similar significant 2.02-fold increase in age-adjusted mutation frequency (*P* = 3.14×10^-8^) when limiting our analysis to donors with matched sperm and blood tissues (**Supplementary Figure 6A**). These observations suggest that mutagenic factors operating independently of age could drive elevated germline mutation in oligozoospermic men.

However, we emphasize that low sperm concentration alone is insufficient to explain the association between subfertility and elevated sperm mutation burdens. Across our longitudinal cohort, which consisted of fertile men whose semen parameters far exceeded the minimum WHO thresholds for normozoospermia at the time of first collection, we analyzed sperm from two donors whose semen collected at the 2nd time point harbored <15 million sperm/mL (**Table 1**). However, these donors were not subfertile in the same regard as men in the oligozoospermic cohort. For one, these donors were part of a therapeutic sperm donation program that required each donor to demonstrate proven fertility such that they never sought fertility care when attempting to conceive a child. Additionally, despite having low sperm concentrations, semen analyses from these donors 2 to 3 weeks after their 2nd collection time point showed that their parameters fell within WHO ranges of normozoospermia (**Supplementary Table 2**), suggesting that the observed reduction in sperm concentration is likely attributed to increased age and variability in semen analyses^72^ rather than acquired male subfertility. When analyzing mutation frequency estimates from these donors, we found that their sperm mutation burdens aligned with measurements from normozoospermic men and were substantially reduced relative to oligozoospermic men (**Figure 3A**). These findings highlight how clinical male infertility, characterized by prolonged difficulties in conception and low sperm concentrations, necessitating fertility care at an andrology clinic, is the key association with germline hypermutation, not simply age-related spermatogenic impairment.

To further characterize germline hypermutation in oligozoospermic men, we next compared age-adjusted mutation frequencies across the custom and control panels, where the custom panel targets coding exons and the control panel tests coding, intergenic, and intronic regions (**Figure 3B**). We performed permutation tests to compare average age-adjusted sperm mutation frequencies (mutation frequency/donor age) from the custom and control panels in normozoospermic and oligozoospermic men (**Methods**). We observed a significant increase in mean mutation frequency in oligozoospermic men across both panels (permutation tests *P* < 0.01), suggesting that increased mutation is likely genome-wide.

Sperm from oligozoospermic men were enriched for C>T transitions, specifically in a non-CpG>TpG context (binomial regression with logit link adjusted *P* = 2.41×10^-15^, **Methods, Supplementary Figure 7**). While non-CpG>TpG transitions are known to be more frequent than transversions in the germline^54^ and may arise through defects in base excision repair^73^, these mutations are signatures of damaged DNA from exogenous sources^74,75^. Therefore, we tested whether a potential enrichment of C>T artifacts may explain the increased sperm mutation frequencies observed in oligozoospermic men. When excluding non-CpG>TpG transitions from the analysis, we still observed a significant 1.34-fold increase in age-adjusted mutation frequencies in oligozoospermic men (**Supplementary Figure 6B**, negative binomial regression *P* = 4.87×10^-3^).

Another potential confounder we addressed is that semen samples are not a pure population of mature gametes. Semen samples contain somatic cells such as leukocytes and epithelial cells from the epididymis^76^ that are expected to have higher mutation rates than gametes. Since sperm concentrations are lower in oligozoospermic men, somatic cells may represent a larger proportion of cells available in the ejaculate of oligozoospermic men relative to normozoospermic men, resulting in artifactually elevated mutation frequencies. To minimize these confounders, we performed a rigorous somatic cell lysis protocol to remove somatic cell contaminants and visually confirmed the absence of detectable somatic cells (**Methods**)^77^. Given our observed sperm and mutation frequencies, our simulations suggest a somatic genome contamination level of >50% is required to explain the 2-fold increase in germline mutation frequencies. At these levels of somatic cell contamination, at least 35,000/70,000 genomes sequenced in a 200ng library would have to be derived from >17,500 diploid somatic cells. This degree of contamination is 6 to 7 times greater than the theoretical maximum number of somatic cells present after performing our somatic cell lysis protocol, where we failed to detect a single somatic cell out of 25 cells analyzed per microliter per semen sample (**Methods**).

### Pathogenic mutation burdens increase with age in both oligozoospermic and normozoospermic men

Altered DNA damage and repair rates during embryonic germline development may underlie our observations of elevated mutagenesis in oligozoospermic men. For example, spermatogonial stem cell (SSC) precursors in oligozoospermic men could undergo increased rates of mitosis when establishing the initial SSC pool capable of spermatogenesis. Under this hypothesis, mutations may accumulate with each cell division such that surviving SSC lineages in oligozoospermic men could exist in a heightened mutagenic state at the onset of puberty. Furthermore, several research efforts have identified genetic factors that stimulate increased rates of germ cell division during embryogenesis^78^ and after puberty^79–81^. Like oncogenesis, gain-of-function mutations in paternal age effect (PAE) genes confer a proliferative advantage for mutant SSCs, resulting in their clonal expansion^82^. While these mutations are subject to positive selection in the male germline, these “selfish mutations”^82^ are often deleterious when transmitted to offspring, as they are implicated in Mendelian diseases such as achondroplasia *(FGFR3*)^83^ and Noonan syndrome (*PTPN11*)^84^.

To determine whether increased gain-of-function and pathogenic (GoF/P) mutation burdens in PAE genes could drive age-independent mutagenesis in oligozoospermic men, we examined mutation burdens in PAE genes that we included in our custom sequencing panel (*FGFR3, FGFR2, PTPN11, HRAS, RET, KRAS, RAF1, BRAF, CBL, SOS1, MAP2K1*, and *MAP2K2*). Based on the extensive evidence of clonal mutation in these genes^79,82,85^, we analyzed all DNMs in PAE genes that were previously implicated in clonal spermatogenesis (“GoF” for Gain of Function, **Supplementary Table 8**) or Mendelian disease (“P” for pathogenic), including those that were flagged by our strict variant filters (**Methods**). Across 473 unique mutations in PAE genes, 27/473 (5.7%) were pathogenic and GoF^79,86,87^, 16/473 (3.4%) were previously linked to a GoF effect, and 76/473 (16.1%) were pathogenic but lacked a known association with clonal spermatogenesis.

To test whether mutations driving clonal spermatogenesis are enriched in oligozoospermic men, we modeled the fraction of GoF/P mutations in PAE genes from normozoospermic and oligozoospermic men using a binomial regression with a logit link (**Methods**). With this approach, and after multiple test corrections using the Benjamini-Hochberg procedure, we found that increased age was significantly associated with an increased fraction of GoF/P mutations in PAE genes in normozoospermic (adjusted binomial regression *P* = 7.79×10^-9^) and oligozoospermic (adjusted binomial regression *P* = 1.35×10^-2^) men, reflecting the expected biology of PAE gene function in facilitating clonal spermatogenesis (**Figure 4A**). After adjusting for age, we found that oligozoospermic men did not harbor a significant increase in GoF/P mutation burdens compared to normozoospermic counterparts (similar y-intercepts, additive binomial regression adjusted *P* = 0.347). Additionally, our model did not report a significant difference in GoF/P DNM accumulation with age (no significant difference between slopes, binomial model with interaction term adjusted *P* = 0.347, **Figure 4A**). These results contradict the mitotically-driven mutagenic model proposed above, which posits that early acquisition of PAE mutations during germline development in oligozoospermic men mediates increased cell division rates and mutagenesis in SSC precursors. Our findings indicate that the accumulation of GoF/P mutations in our currently defined set of PAE is associated with an individual’s age and not their fertility status. Furthermore, in contrast to mutations with likely GoF effects, the fraction of non-GoF/P mutations in PAE genes exhibited no significant relationship with age in normozoospermic and oligozoospermic men (additive binomial regression adjusted *P* = 0.414 and 0.482, respectively), reinforcing our expectation that these mutations are not positively selected for in the male germline (**Figure 4B**).

**Figure 4.**
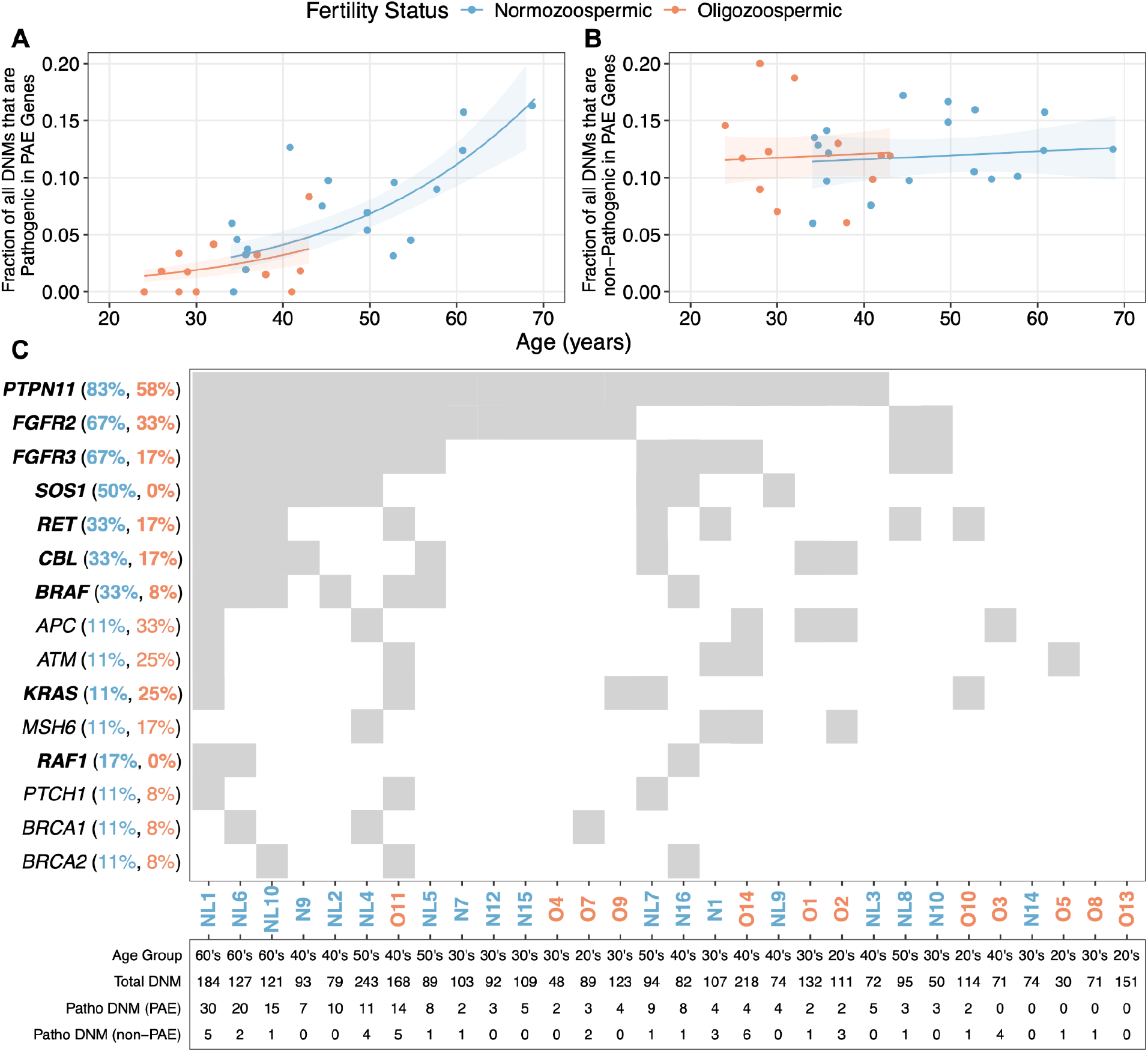
Pathogenic mutations are prevalent in the sperm of normozoospermic and oligozoospermic men. **A)** Fraction of GoF/P mutations in PAE genes are shown for oligozoospermic (orange) and normozoospermic (blue) donors. **B)** Fraction of non-GoF/P mutations in PAE genes. Additive binomial regressions with logit link functions were used to predict GoF/P and non-GoF/P fractions across donor ages. The shaded area represents the 95% confidence interval around these predictions (1.96 times the standard error). **C)** Gene-donor matrix showing the prevalence of ≥1 GoF/P mutation in a given gene. The top 15 genes are sorted in descending order by the total percent of samples with a pathogenic mutation. The percentage of samples within the normozoospermic and oligozoospermic cohorts harboring GoF/P DNMs in each gene is indicated in parentheses. PAE genes are bolded. Donor IDs are included along the x-axis, along with their age at sperm collection, the total number of unique DNMs, unique pathogenic DNMs in PAE genes, and unique pathogenic DNMs in non-PAE genes.

Because GoF/P mutations in PAE genes accumulate with age and result in clonal spermatogenesis, the offspring of older men are at increased risk for inheriting deleterious mutations compared to the offspring of younger men. Consequently, the recommended age cutoff for sperm donation is 40 years old^88^. Across sperm samples analyzed from normozoospermic and oligozoospermic men above and below the age of 40, we recurrently detect GoF/P mutations in PAE genes (**Figure 4C**). These findings mirror prior analyses of sperm and testis samples, which found that donors in their early 20s possess selfish mutations in PAE genes^79,81,89^, suggesting that “reproductive aging” and the accumulation of selfish PAE mutations may commence in young adults well below the age of 40.

While the analysis above focused on gain-of-function and pathogenic mutations in previously characterized PAE genes, our panel may consist of genes that have an undiscovered effect in facilitating clonal spermatogenesis. To examine this possibility, we analyzed pathogenic DNM burdens in non-PAE genes across normozoospermic and oligozoospermic men. Although not statistically significant, likely due to small effect sizes with pathogenic fractions ranging from 0 to 5%, we observed a positive association between increased age and the fraction of pathogenic DNMs in non-PAE genes across normozoospermic (binomial regression adjusted *P* = 0.294) and oligozoospermic men (binomial regression adjusted *P* = 0.152, **Supplementary Figure 8**). While tentative, this finding may indicate that some genes in our custom panel could exert a novel and currently undefined role in promoting SSC proliferation.

Strikingly, we found that oligozoospermic men harbored a significantly increased fraction of pathogenic DNMs in non-PAE genes compared to normozoospermic men (additive binomial regression adjusted *P* = 4.15×10^-2^). Furthermore, we found that *APC, ATM*, and *MSH6* were among the most recurrently mutated genes with pathogenic DNMs across both cohorts (**Figure 4C**). Given that these genes have a well-characterized role in suppressing cellular proliferation and genetic instability in cancer contexts, our findings raise an interesting possibility that inactivating mutations in tumor suppressor genes could also undergo positive selection in the male germline by promoting increased rates of SSC self-renewal, perhaps even in ways that are specific to oligozoospermic men.

### Blood mutation frequencies are not significantly elevated in oligozoospermic men

Epidemiologic studies of infertile men draw an intriguing link between the soma and germline: infertile men and their relatives exhibit poor somatic health relative to normozoospermic men^90–92^. These associations mirror our previously reported findings wherein men in the top quartile of age-adjusted germline mutation rates live five years fewer than men in the lowest quartile^49^. Furthermore, several DNA repair genes implicated in mouse models of male infertility are directly involved in many human cancers^93^. These observations suggest that somatic and germline mutagenesis share a common mechanism in oligozoospermic men, the former manifesting as poor somatic health and the latter as spermatogenic impairment.

On the other hand, existing research highlights a complex discordance between genome mutagenesis, impaired spermatogenesis, and health in infertile men. For example, Lynch Syndrome patients with inherited *MLH1* deficiencies have similar risks for developing male infertility compared to the general population^94,95^, despite the importance of *MLH1* in murine spermatogonial stem cell function. Studies have also shown that azoospermic men with inherited variants in the DNA repair gene, *FANCM*, do not display clinical phenotypes conventionally linked to Fanconi Anemia^96,97^. Additionally, it was previously shown that factors driving elevated mutagenesis exhibit pleiotropy across somatic tissues, and their mutagenic effects are variable across unrelated individuals^98–102^. Indeed, our group showed that unrelated infertile men harbor increased and unique risks for specific blood and non-hematologic cancer types^103^. Recent studies have questioned whether mutation burdens in a specific somatic tissue type, such as blood, could serve as a biomarker for predicting one’s overall health^99,101^. Collectively, these findings further question the relevance of somatic DNM rates to overall health and highlight complexities related to tissue-specific mutagenesis in infertile men.

To test these possibilities, we compared mutation frequencies estimated from duplex sequencing the bulk blood of normozoospermic and oligozoospermic men. At the age of 30, a negative binomial regression model with a log link predicts blood mutation frequencies of 7.68×10^-8^/duplex bp (95% CI: 6.53×10^-8^/duplex bp to 8.82×10^-8^/duplex bp) and 9.73×10^-7^/duplex bp (95% CI: 8.11×10^-8^/duplex bp to 1.13×10^-7^/duplex bp) in normozoospermic and oligozoospermic men, respectively (**Figure 5A**). While our negative binomial model reported that this 26.8% increase in mutation frequency was statistically significant (negative binomial regression *P* = 0.0295), this effect was clearly driven by three oligozoospermic donors (O2, O5, and O12). Indeed, removing any of these three donors from the negative binomial regression analysis yielded a non-significant difference in blood mutation rates (after removing O2 *P* = 0.0650, after removing O5 *P* = 0.0811, after removing O12 *P* = 0.0826). These observations of increased somatic mutagenesis in a subset of infertile oligozoospermic men suggest that blood hypermutation is not a shared feature across all oligozoospermic individuals. Interestingly, these three donors were significantly enriched for CpG>TpG mutations (additive binomial regression adjusted *P* = 7.90×10^-4^), suggesting that their blood genomes may have been subject to increased rates of 5-methylcytosine deamination (**Supplementary Figure 9**).

**Figure 5:**
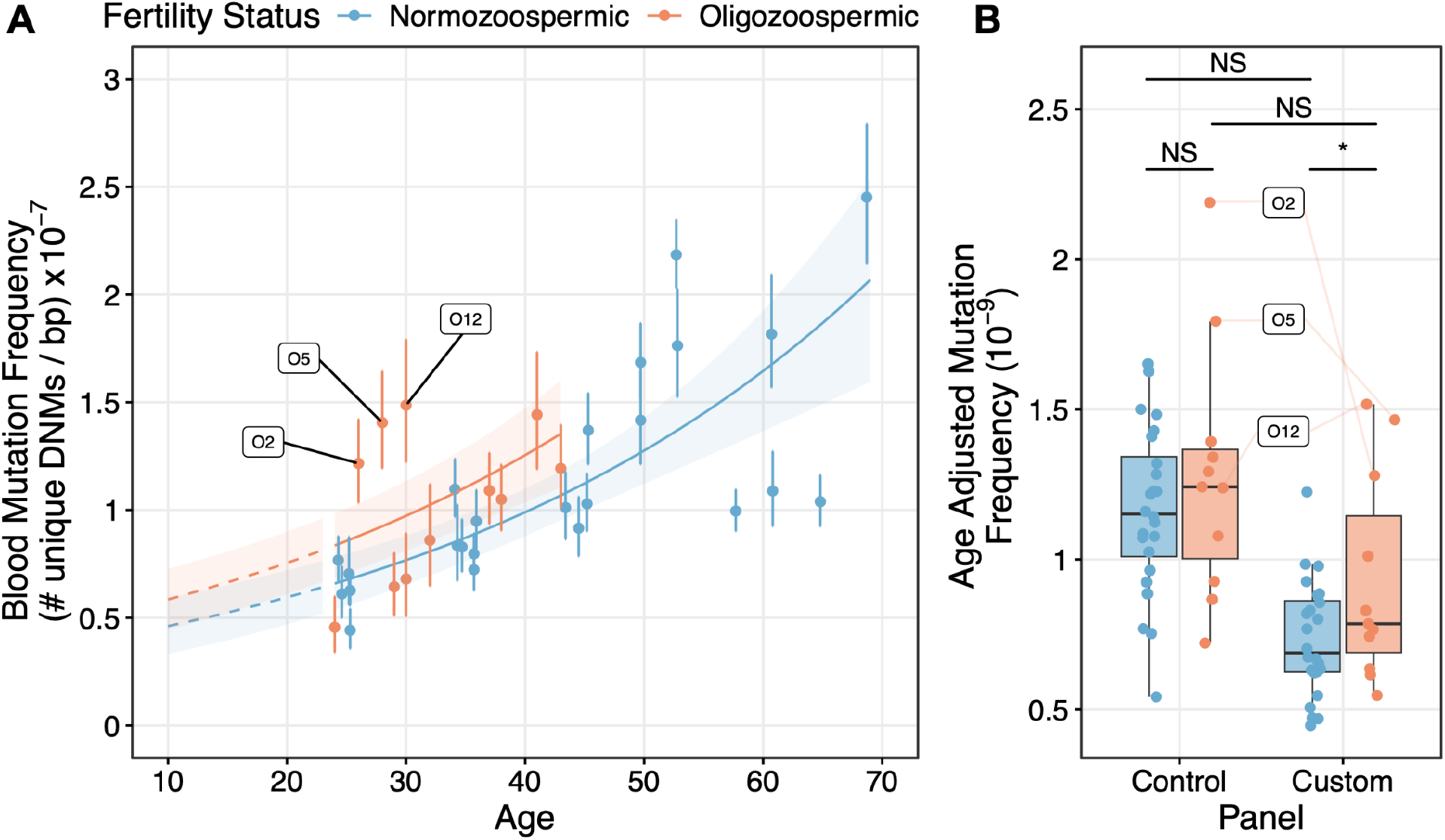
Blood hypermutation is not ubiquitous in oligozoospermic men. **A)** Blood mutation frequencies along with 95% confidence intervals in normozoospermic (blue) and oligozoospermic (orange) donors are shown. Mutation frequency measurements were derived from 200ng non-replicate samples. The standard error from the negative binomial regression model was multiplied by 1.96 to generate the shaded 95% confidence interval. Labeled are three oligozoospermic men whose blood mutation frequency estimate drives our observed significant increase in blood DNM frequencies between oligozoospermic and normozoospermic men. **B)** Age-adjusted mutation frequencies (mutation frequency / donor age) are depicted for each donor when measured from the control and custom panels. NS indicates a permutation test p-value > 0.05. The solid, light orange lines indicate custom and control panel measurements for the three donors labeled in panel A.

Using the same permutation tests described above, we found that age-adjusted blood mutation frequencies in the control panel were not significantly different between normozoospermic and oligozoospermic men (permutation test *P* = 0.1685). However, we observed a significant increase in age-adjusted blood mutation frequencies in the custom panel of oligozoospermic men (permutation test *P* = 0.0245, **Figure 5B**). Similar to the overall blood mutation frequency analysis, however, this custom panel observation was driven by the same three oligozoospermic men described above, where excluding any donor yielded a non-significant difference in custom panel mutation frequencies between normo- and oligozoospermic men (after removing O2 *P* = 0.0548, after removing O5 *P* = 0.0645, after removing O12 *P* = 0.0659). Furthermore, after multiple test corrections on binomial regressions (**Methods**), no gene in the custom panel was found to be significantly enriched for DNMs in the blood of oligozoospermic men, highlighting how somatic mutagenesis in oligozoospermic men is not restricted to a specific gene set but is likely distributed across a variety of coding sequences in the genome.

To determine whether increased sperm mutation frequencies were associated with increased blood mutation frequencies within donors, we examined the correlation between age-adjusted sperm and blood mutation frequencies. However, similar to a previous study that duplex sequenced matched sperm and blood from six 18-year-old males^60^, we failed to identify a significant correlation between age-adjusted sperm and blood mutation frequencies in either normozoospermic (Pearson’s r = −0.1691, *P* = 0.5312) or oligozoospermic men (Pearson’s r = 0.1967, *P* = 0.6119). These findings suggest that sperm mutation frequencies are a poor proxy for predicting blood mutation burdens. Furthermore, our findings indicate that blood mutation frequencies alone are an insufficient biomaker for male infertility and are unable to explain the general epidemiologic association of male infertility with poor overall health.

## Discussion

The etiology of male factor infertility and its association with poor somatic health remains elusive. To investigate potential relationships between genome mutagenesis, reduced sperm production, and poor health in male infertility, we used duplex DNA sequencing to analyze single nucleotide DNM rates and patterns in the sperm and blood of normozoospermic and oligozoospermic men. This inquiry builds upon studies that previously duplex sequenced bulk sperm to study mutations in the paternal age effect gene, *FGFR3*^*61*^, and other loci representative of genome-wide mutagenesis^60,62,63^.

From our bulk sperm analysis, we highlight germline hypermutation as a possible hallmark feature of impaired spermatogenesis, where we observed a significant 1.34 to 2.01-fold increase in age-adjusted mutation frequencies in oligozoospermic men relative to normozoospermic controls. More specifically, our results suggested that this increase is highly associated with a clinical diagnosis of male infertility rather than an age-related reduction in semen parameters. Furthermore, our findings indicated that the male germline in oligozoospermic men is predisposed to hypermutation by the onset of puberty. Our observation of increased germline mutation in oligozoospermic men was additionally robust to several confounding factors, including somatic cell contamination and immotile sperm percentages in a bulk cell population. Recognition of germline hypermutation in infertile men has broad implications for understanding how abnormalities during spermatogonial stem cell self-renewal (mitosis) and differentiation (meiosis) may underlie reduced sperm production and increased germline mutation burdens.

Observing elevated mutation in the sperm of infertile oligozoospermic men demands further understanding of the molecular and cellular mechanisms that contribute to spermatogonial stem cell (SSC) dysfunction and heightened germline mutagenesis. One possibility is that the embryonic germ cell niches in oligozoospermic men may be more mutagenic during development, resulting in increased mutation burdens by the onset of puberty. Building upon the development of the “faulty male hypothesis”^104^, which posits that the male germline is susceptible to mutation accumulation as rates of DNA damage outweigh rates of repair^57^, this would argue that the germline of oligozoospermic men is “faultier.” Perhaps increased oxidative stress^105^, alterations in chromatin remodeling^106,107^, and defects in DNA repair pathways such as cross-link repair^108^ work in tandem to impair cell viability, reduce the number of SSCs available for sperm production at puberty in oligozoospermic men, and increase mutation burdens in surviving germ cell lineages.

Another possibility is that oligozoospermic men possess fewer germ cells during embryogenesis, allowing surviving SSC lineages to rapidly proliferate and occupy the physical space in a developing seminiferous tubule. Current estimates suggest that a given male germ cell lineage will undergo 30 rounds of mitosis following primordial germ cell specification and before puberty onset^109^. This number may be increased in oligozoospermic men, allowing for the accumulation of mutations during more rounds of replication. While we observed recurrent pathogenic DNMs *APC, ATM*, and *MSH6*, raising an intriguing possibility that inactivating tumor suppressor genes could mediate SSC proliferation rates, our analysis of mutations in known PAE genes suggests that PAE-mediate clonal spermatogenesis is less likely to explain increased replication rates in oligozoospermic men. Importantly, under both the replication-independent and dependent models, mutations from early stages of embryonic development would be shared across many spermatogonial stem cell lineages. Should these mutated lineages persist to spermatogenesis, we would expect to see a higher fraction of clonal mutations in the bulk sperm of oligozoospermic men than in normozoospermic men. Furthermore, this early-mutation model suggests that SSCs in oligozoospermic men at puberty would exist at an already elevated mutagenic state.

The final model we have considered is that elevated and age-independent mutagenesis in oligozoospermic men could arise from errors during meiosis. This hypothesis is built from observations in azoospermic men, where meiotically intolerant forms of genomic instability, including aneuploidies, are frequently seen in SSCs and result in their maturation arrest^110,111^. Since meiosis occurs solely in differentiated germ cells, meiotically derived mutations would not be shared with other gametes in a bulk sperm population, resulting in an excess of nonclonal mutations. Furthermore, this “mutagenic spermatogenesis” model implies that oligozoospermic men are predisposed to germline hypermutation as soon as they begin puberty. Future studies are needed to disentangle these mechanistic possibilities, and understanding the underlying mechanism could lead to therapeutic interventions.

We note that our study is limited in that our current duplex sequencing depths prevent us from accurately assessing the true prevalence of a DNM in a bulk sperm population. Consequently, our dataset is ill-equipped to examine the biological validity of pre- and post-pubertal models for mutagenesis. At median duplex depths on the order of 1,000 to 10,000X, we cannot measure the true allele frequency of mutations in semen. This is because even severely oligozoospermic men can possess millions of sperm where clonal mutations shared across multiple sperm cells are likely to be detected from a single gamete at our current duplex depths^62^. To overcome these limitations, even deeper duplex DNA sequencing strategies are needed to assess the true prevalence of mutations in a bulk sperm population. Furthermore, analyses of SSC counts and rates of mitosis/meiosis in the testes of infertile men and animal models across different stages of germline development will illuminate fundamental differences in SSC biology^112–114^. Linking these insights to our observations for increased mutagenesis in oligozoospermic men is necessary to understand the precise mechanisms underlying spermatogenic impairment and develop appropriate treatments for male infertility, which currently does not have a single FDA-approved therapeutic.

Whereas germline hypermutation was observed to be a general feature of oligozoospermia, elevated blood mutation rates were specific to a subset of oligozoospermic men. Although our observations for donor-specificity in blood mutagenesis complicate our understanding of the epidemiologic associations linking male infertility with poor somatic health, several factors can explain our results. We previously analyzed 6,460 pedigrees of non-obstructive azoospermic (NOA), severely oligozoospermic (SO), and fertile men to show that 35% of family members of men with severe male infertility demonstrated an elevated risk for family-specific cancer patterns^103^, raising the question of whether this proportion of men may also harbor increased rates of blood mutagenesis. However, given the family-specific nature of these findings, blood may be a poor proxy for investigating elevated somatic mutagenesis in the context of male infertility, especially if these individuals have elevated risks for developing non-hematologic conditions. Instead, examining somatic mutations in biologically relevant tissues from infertile men stratified by risks for specific somatic comorbidities could uncover individual-specific associations between somatic mutagenesis and poor health. Phenotypic stratification is also essential when considering the pleiotropy of mutation rate modifiers. Numerous studies have demonstrated how inherited defects in DNA repair genes linked to syndromic cancers (*MUTYH*^*101,102*^, *POLE*^*98,99*^, and *POLD1*^*98–100*^) and molecular processes such as gene transcription^115,116^ exert variable effects on mutation rate across tissue types. Indeed, outside of tissues associated with cancer syndromes, elevated somatic mutation burdens are poor predictors of premature aging and additional conditions^99,101^. Therefore, future investigations with large cohorts focused on examining clinically relevant tissues are needed to investigate whether elevated somatic mutation burdens are linked to specific patterns of somatic health in infertile men.

Regardless of whether germline hypermutation occurs before or after puberty, our findings highlight increased germline DNM rates as a hallmark feature of spermatogenic impairment in oligozoospermic men. The clinical utility of this observation is contingent upon future work testing whether sperm mutation profiles can function as predictive biomarkers for assisted reproductive outcomes or personalized therapeutics. To this end, large-scale efforts are needed to comprehensively characterize germline mutagenesis across male infertility subtypes, sperm selection strategies (e.g., swim-up assays), and assisted reproductive technology (ART) procedures (e.g., *in vitro* fertilization). Furthermore, we detected pathogenic mutations in PAE and non-PAE genes in the sperm of normozoospermic and oligozoospermic men below the age of 40. Because these mutations are often found in less than 1 out of 1,000 sperm, these rare mutations are unlikely to be transmitted to the offspring of a given couple^117^. However, over 389,000 ART procedures were conducted in 2022 in the United States^118^, suggesting the overall risk for transmitting low-frequency, pathogenic mutations is non-negligible. Therefore, future improvements to PGT-M (monogenic gene disorders) strategies may include the examination of deleterious variation in PAE genes implicated in congenital disorders. Given the increasing average age of male paternity^119^ and the use of ART worldwide^120^, these advances will inform personalized PGT and ART options that could improve conception and birth rates in infertile couples.

Observing elevated mutation rates in oligozoospermic men with fertility issues motivates future efforts using large cohorts of infertile men spanning a diversity of ages and sperm concentrations to estimate the relationships between germline mutagenesis, fertility status, semen parameters, and increased age. Furthermore, our blood findings suggest that tissue-specific mutagenic mechanisms may underlie distinct patterns of poor somatic health in infertile men and that blood may not be an ideal tissue to predict one’s overall health. Future investigations that stratify infertile men based on shared risks for somatic comorbidities are needed to examine the relationship between poor reproductive and somatic health in male infertility.

## Methods

### Sample acquisition

Bulk sperm and blood samples of normozoospermic and oligozoospermic men were collected from the Subfertility Health Assisted Reproduction and the Environment (SHARE) cohort. Donors from this cohort each provided consent under an institutional review board-approved protocol (#012049) for their tissues to be used for general research use. This tissue repository is a large biobank that, between 2004 and 2011, has accumulated over 5,000 sperm and 2,000 blood from fertile and infertile men between 18 and 89 years of age^43,121^. Semen analyses for individuals in the SHARE cohort were conducted in accordance with the World Health Organization (WHO) standards^4,122^. These analyses measured parameters including sperm concentration (million sperm per mL), sperm count (million), sperm motility (percent of sperm with progressive motility), total motile count (million), and sperm viability (percent of sperm that are viable).

### Cohort description

Ten normozoospermic men comprised the longitudinal cohort. As part of a therapeutic sperm donation program, these donors were required to demonstrate proven fertility, and their semen parameters were all within WHO ranges for normozoospermia. In addition to therapeutic use, these donors consented to their semen being used for general research use under the same IRB above. The semen collected at the time of sperm donation marked the first collection time point. These donors provided an additional semen sample for research use 12 to 24 years after the 1st collection date. The remaining 15 men in the normozoospermic cohort comprised men from male-female couples seeking fertility care at the University of Utah Andrology Clinic. After standard diagnostic workups at the clinic, these individuals were found to have clinically normal semen parameters, including sperm concentration, suggesting that male factor infertility was not a primary driver for the couple’s infertility. In contrast, the oligozoospermic cohort was made up of male andrology patients with abnormally low sperm concentrations, which likely contributed to their infertility case. All normozoospermic and oligozoospermic donors had no history of smoking or chemotherapy exposure.

### Sample DNA preparation

For each semen sample, we performed a somatic cell lysis protocol previously developed by our group^77^. We incubated semen with 0.1% sodium dodecyl sulfate and 0.5% Triton X-100 in diethylprocarbonate water for 20 minutes on ice. Next, we vortexed and aspirated the sample with a 30-gauge needle to mechanically lyse the cells, followed by multiple wash steps to remove DNA and other components of lysed cells. Following this protocol, we confirmed samples were visually free of somatic cells under at least four microscopic fields at 200x magnification on a Makler counting chamber. This verification allowed us to estimate the maximum frequency of somatic cell contamination. For example, a single somatic cell in four microscopic fields on a Makler counting chamber corresponds to a somatic cell concentration of 25,000 cells/ml, or 7500 cells in the final post-somatic cell lysis volume of 300 microliters. With this maximal theoretical somatic cell concentration and the known sperm concentration for each sample, we could estimate the maximum percentage of somatic cell-derived DNA relative to sperm-derived DNA. This estimation was used in the *Somatic cell contamination simulation* described below. DNA extraction for sperm samples following somatic cell lysis was conducted simultaneously using a column-based DNA extraction protocol from the DNeasy kit (Qiagen) modified with cell lysis under reducing conditions. Archived blood DNA across all normozoospermic and oligoozospermic men was extracted from whole blood using the Gentra Puregene Blood Kit according to the manufacturer’s recommendations. Sperm and blood DNA were purified using the Zymo Clean and Concentrator kit. DNA concentration (ng/microliter) was determined using the Qubit dsDNA assay. We prepared sperm libraries from oligozoospermic donors, O13, O7, and O14, by purifying previously extracted DNA. We used 200ng of DNA as input into each duplex sequencing library with selected technical replicate samples using 500ng of DNA. Technical replicates were selected based on having a sufficient amount of extracted DNA available for additional sequencing. Technical sperm replicates from oligozoospermic men were prepared using 200ng of DNA in a separate library preparation and sequencing batch.

### Designing TwinStrand Duplex Sequencing Panel

Conventional Next Generation Sequencing technologies have error rates five orders of magnitude greater than the male germline mutation rate we aimed to measure. This low signal-to-noise ratio renders it very challenging to accurately detect germline mutations with conventional methodologies. To overcome these challenges, we employed duplex sequencing, which overcomes these technical limitations by molecularly barcoding individual DNA fragments and each strand during library preparation. The barcoded amplicons are then sequenced and mapped to the genome to generate a consensus sequence of each strand and then the original double-stranded molecule.

However, because reads from multiple amplicons of each strand are required to confidently distinguish true mutation from error, factors such as sequencing costs and barcode diversity limit the application of deep duplex sequencing to a genome-wide scale. Given these cost constraints, we designed a 325kb targeted panel consisting of exons from 93 genes involved in sperm production, DNA damage repair, cancer, and monogenic forms of male infertility. The full table of genes can be found in **Supplementary Table 1**. Coding exons were included in the panel based on a series of biological criteria described below.

#### Step 1

We selected the 93 genes for inclusion in their panel based on their relevance to spermatogenesis (including PAE genes, **Supplementary Table 8**), male infertility, and somatic health. Here, we included genes implicated in mouse models of infertility and human forms of cancer from Nagirnaja et al., 2018^93^. Additionally, we included genes from a review that identified genes with moderate to definitive association with spermatogenic failure^123^.

#### Step 2

We first identified isoforms annotated as principal isoforms by APPRIS^124^ (2020_11.v37), which designates the “representative isoform” for each gene. Additionally, we used the GTEx dataset (version: June 5, 2017; v8_RSEMv1.3.0) to select isoforms with >1 TPM expression in the testis.

#### Step 3

We targeted functionally relevant exons in each isoform to narrow our panel footprint further. We first identified exons that were annotated with a Pfam functional domain. However, not all isoforms contained exons with Pfam annotations. In these instances, we identified exons corresponding to Uniprot domains. We also included all exons in the isoform cases without known Pfam and Uniprot domains. Finally, we included exons harboring cancer hotspot sites (**Supplementary Table 9**) indicated by three studies^125–127^.

#### Step 4

We designed our targeted sequencing panel to include exons that TwinStrand Biosciences could optimally capture using oligonucleotide probes (Integrated DNA Technologies). Thus, exons corresponding to pseudogenes or those found in low-complexity regions of the genome were excluded.

#### Step 5

Overlapping exons in a given gene were merged using Bedtools^128^ v2.30, resulting in 988 intervals (“custom panel”). We included 20 additional intervals spanning 48kb of noncoding and intergenic loci (“control panel”). The code and set of intervals are provided in the GitHub repository and **Supplementary Table 10**.

### Library preparation and sequencing

Extracted and purified DNA samples (200ng or 500ng) were sheared using an enzymatic fragmentation protocol provided by TwinStrand Biosciences. Compared to mechanical fragmentation with Covaris sonication, this method reduces oxidative damage at the terminal ends of DNA fragments. Fragmented DNA samples underwent library preparation following TwinStrand Biosciences’ Duplex Sequencing Kit Manual ^66,67,129,130^.

With this protocol, DNA fragments are end-repaired and A-tailed with a reaction of 10x NEBNext end-repair buffer and NEBNext end-repair enzyme mix (New England Biolabs). Using a ligation reaction mix (10x Ultrapure ligation buffer, ddH20, Ultrapure T4 ligase, and DuplexSeq adapters), we ligated DuplexSeq adapters to A-tailed fragments. Each DuplexSeq adapter contains the duplex barcode needed to generate a duplex consensus sequence (described below) and an Illumina adapter required for sequencing. Next, we conditioned the library with a mixture of DNA repair enzymes (TwinStrand) to reduce the damage introduced during library preparation. These barcoded DNA products were then size selected with SPRIselect beads and PCR amplified with the Bio-Rad T100 thermal cycler. This PCR step generated multiple DNA copies from the original template fragment, including the original duplex barcode. After adding SPRIselect beads to select amplified products with desired lengths between 300bp to 600bp, we performed the first round of probe hybridization using biotinylated ssRNA probes that, upon the addition of streptavidin-beads, captured DNA fragments corresponding to desired genomic regions from our custom and control panels. We executed these PCR, size selection, and capture steps a second time to further maximize on-target DNA yield. Finally, after a third round of PCR, we added SPRIselect beads to enrich for amplified products with desired fragment lengths.

The DNA concentration in each prepared library was then quantified with Qubit dsDNA assay. Libraries with >7.69ng/uL and 19.2 ng/uL in 200ng and 500ng libraries, respectively, were sent to the University of Utah Health Sciences Center DNA Sequencing Core for further quality control assessment using Agilent TapeStation and sequencing. Duplex sequencing libraries were paired-end sequenced using a NovaSeq 6000 machine with 151 base pair reads. Tapestation analysis identified overly fragmented libraries where a large proportion of fragments had lengths <300bp.

### Using Peak Tag Family Size to modulate the number of clusters needed for each library

Duplex sequencing molecularly barcodes individual DNA fragments that are then amplified and sequenced. Reads with matching barcode sequences are part of a “family” derived from the same original DNA molecule. Because of the non-complementary nature of the Illumina adapter sequences that are part of the DuplexSeq barcode, it is possible to delineate whether a read corresponds to the top or bottom strand of the original DNA molecule. Therefore, all fragments in a final library will possess a “family size” corresponding to the number of reads derived from the top and bottom strands of the original DNA molecule. A sufficiently large family size is needed to generate a duplex consensus sequence. This is because the consensus sequence leverages information from the top and bottom strands to represent complementary bases (and mutations) in a double-stranded DNA molecule and generate a “duplex read.” Furthermore, this approach helps resolve PCR or sequencing errors that manifest on amplicons from either strand of the original template molecule.

Because “family size” quantifies the number of amplicons derived from an original DNA molecule, a duplex sequencing library will possess a distribution of family sizes. Here, “peak tag family size” is a qualitative metric that describes the distribution of “family sizes” in a given library. Specifically, PTFS refers to the family size bin that contains the largest proportion of duplex reads in a library. We followed recommendations from TwinStrand Biosciences to target a PTFS of ≥7 for each duplex sequencing library^66^, as this would maximize the number of DNA molecules with top and bottom strand read support and without generating redundant sequencing reads that do not contribute to the generation of a consensus double-stranded DNA molecule. Put another way, when PTFS is low, mean duplex depth will be low as few molecules have sufficient read support to generate a duplex consensus. As PTFS increases, more molecules will reach consensus and generate a duplex read. Eventually, most molecules will have reached a consensus, such that a library’s mean duplex depth becomes saturated.

### Assessing duplex sequencing library complexity

Depending on the complexity of a library, two libraries may possess variable mean duplex depth values when a PTFS ≥7 is reached. For example, a 200ng sample has fewer DNA molecules (lower molecular complexity) than a 500ng sample (higher molecular complexity). As per recommendations from TwinStrand Biosciences, given the size of our custom and control panels, we would anticipate that ∼1 billion and ∼2.5 billion raw reads are required to adequately sequence a 200ng and 500ng duplex sequencing library, respectively. However, low-complexity libraries with a reduced quantity of unique DNA molecules could be caused by errors during library preparation, such as failed probe hybridization. Consequently, these libraries would require fewer reads to reach duplex depth saturation. Therefore, we excluded low-complexity libraries from downstream analyses (except for use during mutation filtering; see below). Specifically, we excluded sperm and blood libraries from normozoospermic and oligozoospermic men that achieved mean duplex depths <2000x even after reaching a PTFS ≥7 (**Supplementary Figure 10**). Additionally, we excluded donor O14’s blood, as this was an overly complex library that reached a mean duplex depth of ∼24000x despite having a PTFS of 4.

### Identifying samples for exclusion

We additionally used a Jaccard statistic to exclude samples showing evidence of cross-contamination. Here, we subset our mutation dataset to ostensibly inherited variants (allele frequency > 0.2, **Equation 1**). Sperm and blood samples from the same donor exhibited Jaccard-statistic values > 0.96. We excluded samples that harbored Jaccard values of >0.7 when examining unrelated donors. This analysis identified samples from N4 (blood) and NL2 (sperm from the second time point) (**Supplementary Figure 11**) as subjects of cross-contamination, warranting their exclusion from downstream analyses. However, we used these samples in our mutation filtering pipeline to flag recurrent mutations detected across multiple sequencing libraries (see “Variant Calling from Duplex BAM”).

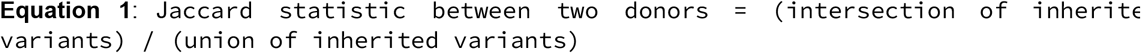

We used the median absolute deviation base function in R to exclude samples whose age-adjusted mutation frequency was greater than 3.5 * MAD relative to non-replicate samples in a given fertility (normozoospermic or oligozoospermic n) and tissue (sperm or blood) group (**Supplementary Figure 12**). Age-adjusted mutation frequencies were calculated by dividing a sample’s mutation count by the donor age. Samples identified as outliers by the MAD test exhibited a significant enrichment for C>A and C>T (non-CpG context), which have been linked to artifacts from library preparation and sequencing^131,132^. Similar to above, however, these samples were used to flag recurrent mutations in our duplex datasets (see “Variant Calling from Duplex BAM”).

### Fastq to Duplex BAM

We developed a custom script to merge sample FASTQ files from multiple S4 flow cell lanes. We then used the nextflow nf-core/fastquorum workflow (version 23.10.1) ^133^ that leverages Fulcrum Genomics’ fgbio duplex sequencing pipeline (version 2.0.2) to align reads from each paired FASTQ file to the Human Reference Genome (GRCh38) and generates a duplex BAM file (also uses fastqc version 0.11.09, multiqc version 1.11). The code and parameters used for running the nf-core/fastquorum workflow are provided in the below GitHub repository. The parameters we explicitly specified are described below.

| Step | Parameter | Justification/Interpretation |
| --- | --- | --- |
| nextflow run | -r aab72e6473 | Use specific fastquorum commit |
|  | --mode 'ht' | High-throughput data processing |
| ExtractUmisFromBam | --read-structure 8M1S+T | 8nt barcode + 1 spacer nucleotide + template DNA |
| GroupReadsByUMI | --groupreadsbyumi_edits 1 | Allow for 1 mismatch in UMIs belonging to the same original DNA molecule (family) |
| CallDuplexConsensusReads | --call_min_reads '3 3 3' | Requires an overall minimum family size of 6 with 3 single strand consensus reads of support from each strand to generate a duplex consensus for a given base |
|  | --call_min_baseq 20 | Ignores bases in reads from the FASTQ file with a PhredScore < 20 |
| FilterConsensusReads | --filter_min_reads '3 3 3' | Bases in duplex consensus reads with <3 reads of single strand support from either strand are masked to N's |
|  | --filter_min_baseq 45 | Bases on a duplex read with a Phred score < 45 are replaced with 'N' |
|  | --filter_max_read_error_rate 0.2 | Remove duplex reads with an error rate > 20% |

### Variant Calling from Duplex BAM

Variant calling was done on duplex BAM files with VarDictJava using the below workflow (https://github.com/AstraZeneca-NGS/VarDictJava)^134^ to generate a VCF. We further annotated each variant with VEP^135^ (v104) and ClinVar^136^ pathogenicity annotations (March 28, 2022). Additionally, we identified genes and variants with likely paternal age effects (PAE) based on prior literature^79,82,85^. We further developed a tool called fraguracy^137^ to identify discrepant nucleotide calls in overlapping portions of paired duplex consensus reads from a sample’s duplex BAM file. Because read pairs are derived from the same original DNA molecule, the sequence in overlapping regions should match exactly, while mismatches may arise at error-prone sites in the genome.

We further imposed a strict set of filters to identify a high-quality list of mutations.

1. Mutation passes VarDict filters (listed below)
2. Mutation does not overlap clonal multi-nucleotide variants (MNV)
3. Mutation not within ten bases of a clonal insertion or deletion (INDEL)
4. Mutation calls on read with < 5% of N calls
5. Mutation at site with < 5% N frequency
6. Mutation not within a homopolymer sequence
7. Mutation not at sites identified by fraguracy (https://github.com/brentp/fraguracy) to harbor > 3 errors

Additionally, we removed mutations based on different levels of recurrence. To identify *de novo* mutations, we performed germline filtering to remove any mutations in the sperm and blood from a given donor, regardless of their allele frequency. Next, we aggregated all sperm and blood mutations across all normozoospermic and oligozoospermic donors and removed mutations found in the opposite tissue. For example, we filtered out sperm mutations from a normozoospermic donor that were found in the donor’s blood sample (within donor recurrence), any blood sample from another normozoospermic (within cohort recurrence) or oligozoospermic (across cohort recurrence) donor. We did not employ a within-tissue recurrence filter (e.g., flagging mutations found in multiple sperm samples) because we expect some mutations, especially those in paternal age effect genes, to be recurrently sperm-specific as they confer a proliferative advantage in spermatogonial stem cells.

| Vardict Step | Parameter | Justification/Interpretation |
| --- | --- | --- |
| VardictJava | -p | Performs nucleotide pileup regardless of the frequency |
|  | -K | Includes unknown nucleotide calls (N's) when reporting total depth at a given site |
|  | -k 1 | Performs local realignment |
|  | -r 0 | Minimum number of reads needed to support a variant; otherwise the variant call is flagged. Setting r to 0 allows for the inclusion of singleton calls supported by a single duplex read |
|  | -p 0 | Calls variants at all positions along a read |
| teststrandbias.R | Default parameters | N/A |
| var2vcf.pl | -A | Outputs all variants called by VardictJava at the same position |
|  | -E | Do not print the END tag |
|  | -f 0.0 | Does not flag any variants for having too low of an allele frequency (does not flag singleton calls) |
|  | -v 1 | Does not flag any variants for having too low of an alternate allele count (does not flag singleton calls) |
|  | -P 0 | Set to 0 to not flag variants based on read position |
|  | -p 1 | Includes variants called along all positions of a read |
|  | -d 3 | Requires a minimum total depth of 3 (this is the default) |
|  | -Q 10.0 | Flags variants with mapping quality < 10, except for variants with allele frequencies > 0.8 |
|  | -m 8.0 | Flags variants if the variant is called on a read with > 8 mismatches |

### Calculating mutation frequency

With the mutations that passed filters, we then calculated mutation frequencies for each sample using **Equation 2**. Because we used an array of filters to identify a more high-confidence set of mutations, we employed the same set of filters (described above) to determine a high-confidence set of bases from which we could detect mutations. To do this, we developed a tool called pbr (v.0.1.6)^138^, which applies nucleotide masking (N calls) and site-based filters to a duplex BAM file. We additionally ignored bases at sites where a cross-tissue recurrent mutation was identified. With this output, we could calculate the total number of high-quality bases sequenced in a sample to measure the total duplex depth achieved at each site.

To calculate sample mutation frequencies, we divided the number of unique DNMs detected in a sample by the total number of high-quality duplex bases sequenced (**Supplementary Tables 6 and 7**). This approach avoids double-counting clonal mutations found across multiple cells, which are more likely to have originated from a single mutational event in a progenitor stem cell that underwent clonal expansion.

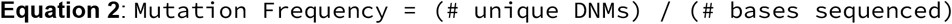

#### pbr command with filters

~~~
$pbr --threads 8 --fasta $fasta $bam_file \
       “return string_count(read.sequence, ‘N’) < 0.05 * read.length and
read.distance_from_5prime >= 15 and read.distance_from_3prime >= 15 and
bit32.band(read.flags, bit32.bor(4, 256, 512, 1024)) == 0” \
       --max-depth 100000 \
       -p “return pile.n / pile.depth < 0.05” \
       --bedfile $subset_intervals
~~~

#### pbr command without filters

~~~
$pbr --threads 8 --fasta $fasta $bam_file \
      “return true” \
      --max-depth 100000
      --bedfile $all_intervals
~~~

### Confidence intervals around mutation frequency

Given a sample’s mutation count, we estimated the 95% confidence interval around the Poisson mean using Equations 3a and 3b. The upper and lower mutation counts measured from these equations were divided by total duplex bases sequenced to generate 95% confidence intervals around each sample’s mutation frequency estimate.

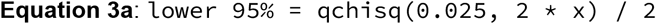

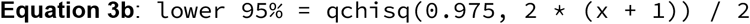

### Downsampling BAM files

After filtering out bases called at error-prone sites and low-quality reads, the 200ng libraries of sperm and blood from normozoospermic men had ∼2,500x deeper duplex depth than the tissues of oligozoospermic men (**Supplementary Figure 1**). We evaluated mutation frequency estimates from downsampled sequencing BAM files from normozoospermic men to rule out the effects of discrepancy median duplex depths when comparing mutation frequencies across cohorts. Normozoospermic blood samples had median duplex depths of ∼6,500x, while oligozoospermic blood samples had median duplex depths of ∼3,500x. As such, blood BAM files were downsampled by 50% using samtools (version 1.18). Similarly, normozoospermic and oligozoospermic sperm samples had median duplex depths of ∼6,250x and 4,000x, respectively. As such, sperm BAM files were downsampled by 33%. We used the pipelines above to perform variant calling, filtering, and mutation frequency calculations. These downsampled values were compared to mutation frequency estimates without downsampling (**Supplementary Figure 4**).

### Examining consistency in mutation frequency estimations without matched tissues

Due to the sampling filtering steps described above (library complexity, Jaccard statistic, MAD outlier test), we analyzed DNM rates and patterns in only one tissue type (either sperm or blood) for some donors (**Table 1**). While failed libraries were still used to flag and remove recurrent DNM within and across individuals, it is possible that having a successfully prepared matched tissue could influence mutation frequency estimates. To demonstrate the consistency in mutation frequency calculations with and without a matched tissue, we first identified donors with successfully prepared sperm and blood libraries with 200ng of DNA. We then measured mutation frequencies by applying all the recurrence filters described above (within donor, within cohort, and across cohort recurrence). Next, we calculated mutation frequencies without applying the within-donor recurrent filters and compared the values derived from these two approaches (**Supplementary Figure 5**). The substantial overlap in 95% confidence intervals between mutation frequencies estimated with and without a matched tissue suggests that our current set of filters can capture DNMs that would be flagged as within-donor recurrent.

### Permutation test to compare sperm and blood custom vs. control panel mutation frequencies

We compared age-adjusted mutation frequencies in sperm and blood by dividing the observed mutation frequency in each donor by the donor’s age. Next, we ran 10,000 permutation tests that randomly restructured the data groupings for age-adjusted mutation frequency assignments. For example, to compare age-adjusted mutation frequencies measured from the custom and control panel in the sperm of oligozoospermic men, we randomly assigned each value with a “custom panel” or “control panel” designation. We calculated the mean difference between each group with these randomized assignments. The distribution of this test statistic was used to generate a p-value equal to the fraction of permutation tests showing a mean difference greater than the/ observed mean difference between groups.

### Estimate DNM accumulation rates in the longitudinal cohort

To calculate DNM accumulation rates across longitudinally collected samples in normozoospermic men comprising the longitudinal cohort, we first restricted our analysis to donors with sperm samples with successfully prepared libraries at both time points. For donors with 200ng and 500ng replicates at a given time point, we calculated an overall mutation frequency estimate by dividing the unique number of DNMs found across both replicates by the sum of duplex bases sequenced across both replicates. We then used the below equations to estimate the DNM accumulation rate per year per 3.1 billion bases.

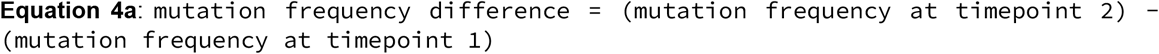

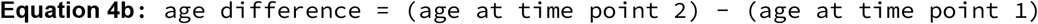

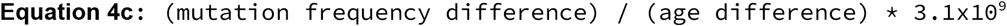

### Mutation frequency regressions in the longitudinal cohort

To model age and donor effects on overall mutation count (n_var) estimates in the longitudinal cohort, we devised a Poisson regression model in R using the donor’s age. Additionally, we included an offset term with a log link function that directly accounts for variation in duplex depth across samples.

#### Model 1 - without Donor ID

~~~
glm(n_var ∼ age + offset(log(depth), family=poisson(link=“log”))
~~~

We next examined how donor-specific age effects could impact mutation burdens by adding the interaction term below.

#### Model 2 - with Donor ID

~~~
glm(n_var ∼ age * donor_id + offset(log(depth), family=poisson(link=“log”))
~~~

The fit between these two poisson models (with and without donor_id interaction term) was compared using an analysis of variance (ANOVA) test. This test reported that a model that accounts for variation in age effects across donors significantly fits the data better than a model agnostic to donor ID (*P* = 5.078×10^-79^).

~~~
anova(Model1, Model2, test = “Chisq”)
~~~

### Overdispersion test for Poisson regressions

To test for overdispersion in our dataset, we used the dispersiontest function from the AER package^139^ in R. When tested on the best-fitting model for the longitudinal sperm analysis (Model 2 above), this test did not report overdispersion, suggesting that a Poisson regression could appropriately model longitudinal sperm mutation frequencies across ages and donors.

~~~
model = glm(n_var ∼ age * donor_id + offset(log(depth),
family=poisson(link=“log”))
AER::dispersiontest(model)
~~~

### Sperm and blood mutation frequency regressions in normozoospermic and oligozoospermic men

To identify factors that impact sperm and blood mutation frequencies, we generated similar Poisson regression models as described above. For this analysis, we included a fertility_status term to compare differences in mutation burden across normozoospermic and oligozoospermic men. When analyzing sperm mutation frequencies, we included an avgMotil term that accounts for differences in the percentage of motile sperm in each semen sample. These analyses were limited to sperm and blood samples prepared from 200ng DNA libraries and excluded replicate libraries.

~~~
glm(n_var ∼ age + fertility_status + avgMotil + offset(log(depth), family = poisson(link = “log”))
~~~

However, we found that in both sperm (*P* = 0.003068) and blood (*P* = 0.002923) using the AER::dispersiontest, the Poisson assumption of equidispersion (variance = mean) was violated as the data was overdispersed (variance> mean). To account for this, we modeled sperm and blood mutation using negative binomial regressions (with the glm.nb function from MASS package^140^ in R) that employed a log link and an offset term to account for differences in duplex depths across samples. Furthermore, these negative binomial regressions were used to model sperm mutation frequency estimates from normozoospermic and oligozoospermic men without C>T (non-CpG context) mutations as well as situations where we subsetted our mutation datasets to donors with successfully prepped matching sperm and blood samples.

~~~
MASS::glm.nb(n_var ∼ age + fertility_status + avgMotil + offset(log(depth), link = “log”)
~~~

In addition to the additive negative binomial regression model, we tested whether a significant interaction exists between age and fertility status. To this end, we ran the below regression and compared its fit to an additive regression model using AIC scores. Additionally, we used an ANOVA test to determine if including the interaction term significantly improved upon the additive model’s fit to the data.

~~~
MASS::glm.nb(n_var ∼ age * fertility_status + avgMotil + offset(log(depth), link = “log”)
~~~

To examine age effects in a cohort-specific manner, we subsetted the data to mutation frequencies estimated from either the normozoospermic or oligozoospermic cohort and removed the fertility status term from the negative binomial regression formula, as shown below.

~~~
MASS::glm.nb(n_var ∼ age + avgMotil + offset(log(depth), link = “log”, data = mutation_frequencies[fertility_status == “Normozoospermic”])
~~~

### Mutation spectra regressions in normozoospermic and oligozoospermic men

To examine differences in mutation spectra across normozoospermic and oligozoospermic men, we calculated the fraction of DNMs falling with seven mutation classes: C>A, C>G, C>T (non-CpG), CpG>TpG, T>A, T>C, and T>G (**Equation 5**). We then developed a binomial regression model with a logit link to investigate the effects of age and fertility status on each mutation class fraction. To account for differences in the number of mutations detected across samples, we weighted each sample by the total number of DNMs. When modeling the mutation spectra in sperm, we included avgMotil as a covariate. This term was excluded when analyzing mutation subtype fractions in blood.

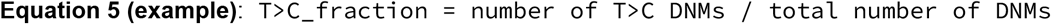

~~~
glm(T>C_fraction ∼ age + fertility_status + avgMotil, family = binomial(link = “logit”), weights = model_df$n_var)
~~~

To independently model age effects in normozoospermic and oligozoospermic cohorts, we subset the mutation fraction dataset to the cohort of interest and removed the fertility status term from the binomial regression formula.

~~~
normo_df = model_df[fertility_status == “Normozoospermic”]
glm(T>C_fraction ∼ age + avgMotil, family = binomial(link = “logit”), weights = normo_df$n_var)
oligo_df = model_df[fertility_status == “Oligozoospermic”]
glm(T>C_fraction ∼ age + avgMotil, family = binomial(link = “logit”), weights = oligo_df$n_var)
~~~

We ran the three regression models described above on each mutation subtype. We then employed a Benjamini-Hochberg multiple-test correction to generate a set of adjusted p-values (**Supplementary Figures 7 and 9**).

### DNM fractional regressions in normozoospermic and oligozoospermic men

To perform our pathogenic fraction mutation analysis, we first calculated the fraction of gain-of-function/pathogenic (GoF/P) mutations in paternal age effect (PAE) and pathogenic mutations in non-PAE genes (**Equation 6**). Gain-of-function mutations were those in PAE genes that were previously shown to be associated with clonal spermatogenesis^79,82,85^ (**Supplementary Table 8**). Pathogenic mutations were annotated from ClinVar (March 28, 2022) with the following terms: pathogenic, pathogenic/likely pathogenic, and likely pathogenic. For this analysis, we also recovered mutations with known gain-of-function and pathogenic effects in PAE genes that were flagged by our strict filters. This adjustment recovered mutations such as chr4:1804392G>A in *FGFR3*, whose position is within a homopolymer sequence and was annotated as a potential error. We then fit a binomial regression model with a logit link function described above to model GoF/P and P fractions in PAE and non-PAE genes using age, fertility status, and sperm motility as covariates. We independently examined age effects in normozoospermic and oligozoospermic men by subsetting the pathogenic DNM fraction dataset to the appropriate cohort. The Benjamini-Hochberg procedure was used to reduce our false discovery rate and generate a set of adjusted p-values. This analysis was limited to 200ng sperm samples and excluded replicate libraries.

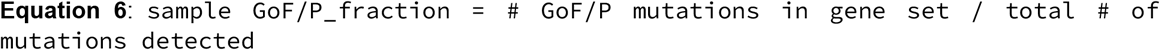

~~~
glm(GoF/P_fraction ∼ age + fertility_status + avgMotil, family = binomial(link = “logit”), weights = model_df$n_var)

normo_df = model_df[fertility_status == “Normozoospermic”]
glm(GoF/P_fraction ∼ age + fertility_status + avgMotil, family = binomial(link = “logit”), weights = normo_df$n_var)

oligo_df = model_df[fertility_status == “Oligozoospermic”]
glm(GoF/P_fraction ∼ age + fertility_status + avgMotil, family = binomial(link = “logit”), weights = oligo_df$n_var)
~~~

This same approach was used to model the potential enrichment of blood-derived DNMs in oligozoospermic men across individual genes in the custom panel.

### Generating predictions from regression models

To generate the regression curves (predicted mutation frequency and 95% confidence intervals) shown in Figures 3, 4, and 5, we used the predict function in R with the appropriate model as its input. We used an ANOVA test in each case to determine whether a nested additive or an interaction model better fit the data in question (**Supplementary Table 11**). The better fitting model was used to generate the predicted datasets and plots to depict each dataset graphically (see GitHub link).

### Somatic cell contamination simulation

Rather than possessing a homogenous population of sperm cells, an individual’s ejaculate contains somatic cells such as leukocytes and epididymal epithelial cells. This reality could confound our observations, where assuming there is an equal number of somatic cells in a normozoospermic and oligozoospermic male’s ejaculate sample, the fraction of cells that are somatic in origin is increased in oligozoospermic men. We performed the following simulation to determine whether an enrichment of somatic cell contamination could explain the elevated mutation frequencies in subfertile men. A streamlit application for running the simulation can be found in the provided GitHub repository.

1. Assume each normozoospermic and oligozoospermic sample uses 70,000 cells as input for a 200ng library.
2. Set normozoospermic and oligozoospermic germline mutation frequencies to be equivalent. This value is based on observed sperm mutation frequencies in normozoospermic men. Here, we assume that oligozoospermic and normozoospermic germline mutation frequencies are equivalent. Germline mutation frequency @ 30 y/o: oligozoospermic = 3.41×10^-8^; normozoospermic = 3.41×10^-8^
3. Similarly define equivalent normozoospermic and oligozoospermic somatic mutation frequencies. This value is based on blood mutation frequency estimates in oligozoospermic men. Observed somatic mutation frequency @ 30 y/o: oligozoospermic = 9.73×10^-8^; normozoospermic = 9.73×10^-8^
4. Set the baseline degree of somatic genome contamination in fertile men. We set this at 1%
5. Iterate over 1% to 500% somatic genome contamination in oligozoospermic men.
6. During each iteration, calculate the aggregated mutation frequency from somatic and germline genomes in normozoospermic and oligozoospermic men.
7. Determine the somatic cell contamination % in subfertile men at which the ratio between (subfertile mutation frequency) / (fertile mutation frequency) > 2

## Supporting information

Supplementary Manuscript

Supplementary Table 1 - Gene list

Supplementary Table 2 - Longitudinal donors

Supplementary Table 3 - Duplex bases removed

Supplementary Table 4 - Mutations removed

Supplementary Table 5 - Mutation dataset

Supplementary Table 6 - Sample coverage dataset

Supplementary Table 7 - Sample interval coverage dataset

Supplementary Table 8 - PAE var list

Supplementary Table 9 - Cancer hotspot list

Supplementary Table 10 - Probe intervals

Supplementary Table 11 - Interaction vs additive models

Supplementary Figure 1 - Sample coverage

Supplementary Figure 2 - Nonclonal and clonal DNM fraction

Supplementary Figure 3 - Mutation frequency in technical replicates

Supplementary Figure 4 - Downsampled normozoospermic BAM mutation frequencies

Supplementary Figure 5 - Matched donor vs recurrence filter mutation frequencies

Supplementary Figure 6 - Matched donor and non-C>T mutation frequencies

Supplementary Figure 7 - Sperm mutation spectra

Supplementary Figure 8 - Fraction pathogenic in non-PAE genes

Supplementary Figure 9 - Blood CpG>TpG spectra

Supplementary Figure 10 - Library complexity

Supplementary Figure 11 - Jaccard

Supplementary Figure 12 - MAD mutation frequency outliers

## Data Availability

All data produced and code are available online at https://github.com/quinlan-lab/normo-vs-oligo-manuscript or in the supplementary material. In the interest of patient privacy, we are unable to provide precise donor ages in our publicly available dataset but are able to provide such data upon reasonable request.

## Code availability

The code to conduct all of the analyses can be found here: https://github.com/quinlan-lab/normo-vs-oligo-manuscript

