## Supplementary Manuscript for "Sperm from infertile, oligozoospermic men have elevated mutation rates"

**Supplementary Figures**

Supplementary Figure 1 - Sample coverage
Supplementary Figure 2 - Nonclonal and clonal DNM fraction
Supplementary Figure 3 - Mutation frequency in technical replicates
Supplementary Figure 4 - Downsampled normozoospermic BAM mutation frequencies
Supplementary Figure 5 - Matched donor vs recurrence filter mutation frequencies
Supplementary Figure 6 - Matched donor and non-C>T mutation frequencies
Supplementary Figure 7 - Sperm mutation spectra
Supplementary Figure 8 - Fraction pathogenic in non-PAE genes
Supplementary Figure 9 - Blood CpG>TpG spectra
Supplementary Figure 10 - Library complexity
Supplementary Figure 11 - Jaccard
Supplementary Figure 12 - MAD mutation frequency outliers

**Supplementary Tables**

Supplementary Table 1 - Gene list
Supplementary Table 2 - Longitudinal donors
Supplementary Table 3 - Duplex bases removed
Supplementary Table 4 - Mutations removed
Supplementary Table 5 - Mutation dataset
Supplementary Table 6 - Sample coverage dataset
Supplementary Table 7 - Sample interval coverage dataset
Supplementary Table 8 - PAE var list
Supplementary Table 9 - Cancer hotspot list
Supplementary Table 10 - Probe intervals
Supplementary Table 11 - Interaction vs additive models

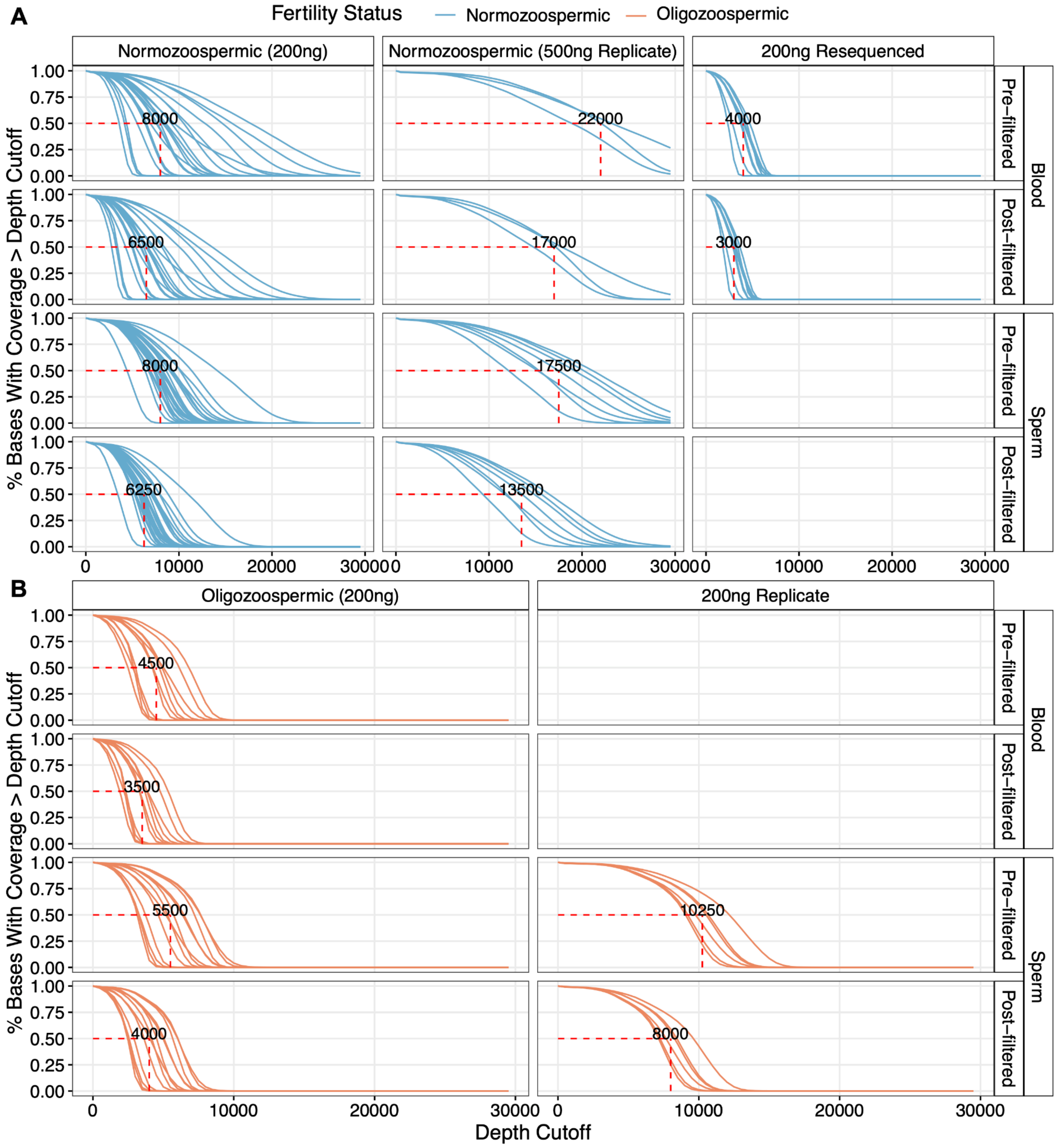


**Supplementary Figure 1**: Each curve depicts the fraction of bases (y-axis) covered at a given duplex depth indicated along the x-axis for a given sample. Sample libraries are split based on the input DNA amount, tissue, pre/post filter for duplex coverage (**Methods**), and fertility status. Libraries derived from (**A**) normozoospermic and (**B**) oligozoospermic men are colored blue and orange, respectively. Only samples with successfully prepared libraries (**Methods)** are depicted. These filters were similarly applied to mutation calls. The red dashed line indicates the median duplex depth at which 50% of bases are covered across all libraries per group.

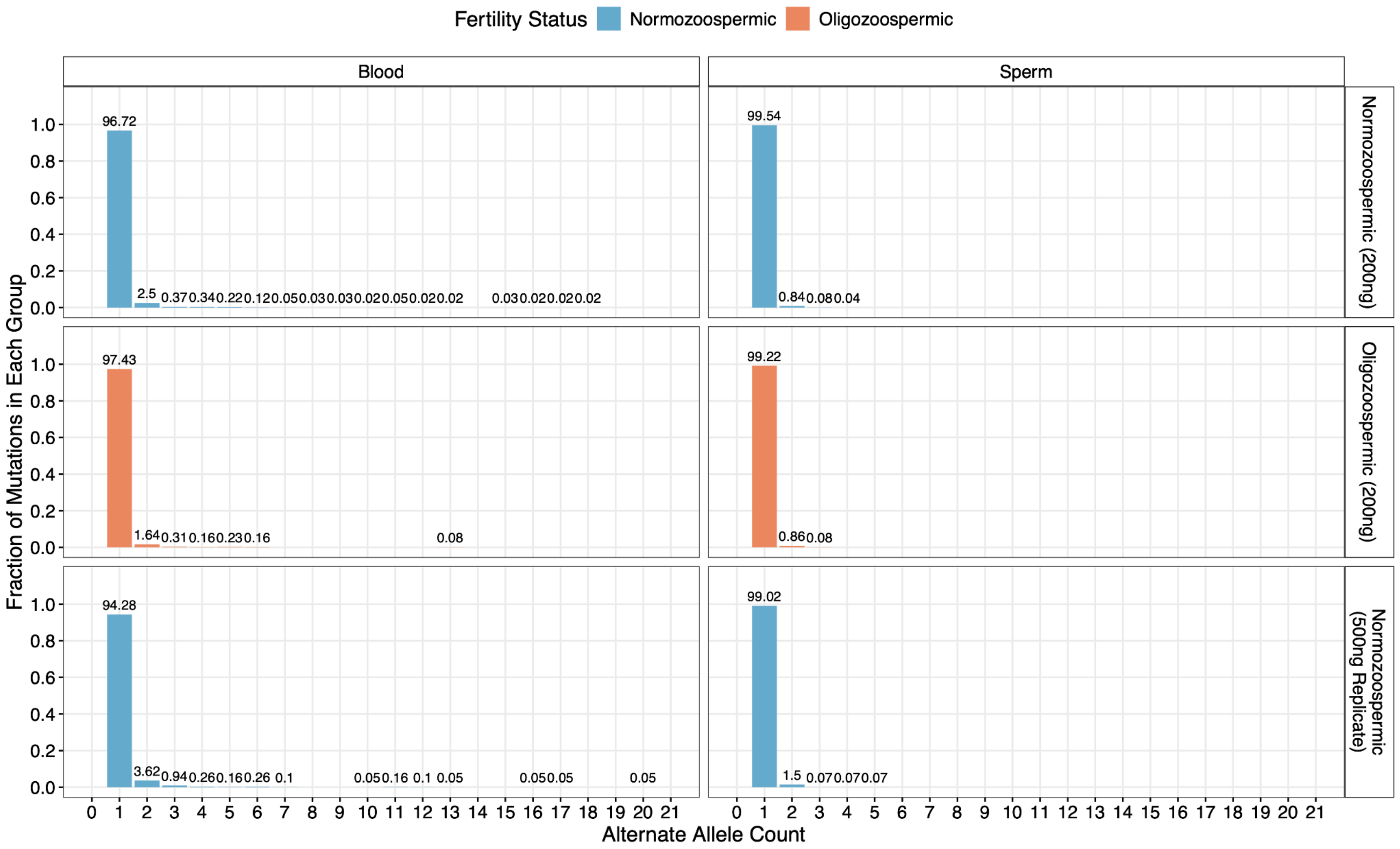


**Supplementary Figure 2**: The fraction of post-filtered DNMs are binned by their alternate allele count across all samples in each fertility status and input DNA group (row-wise) and tissue (column-wise). Clonal mutations had an alternate allele count >2, as they were detected in at least two duplex reads. Each bar depicts the percentage of mutations within each group that fall into the corresponding alternate allele count bin.

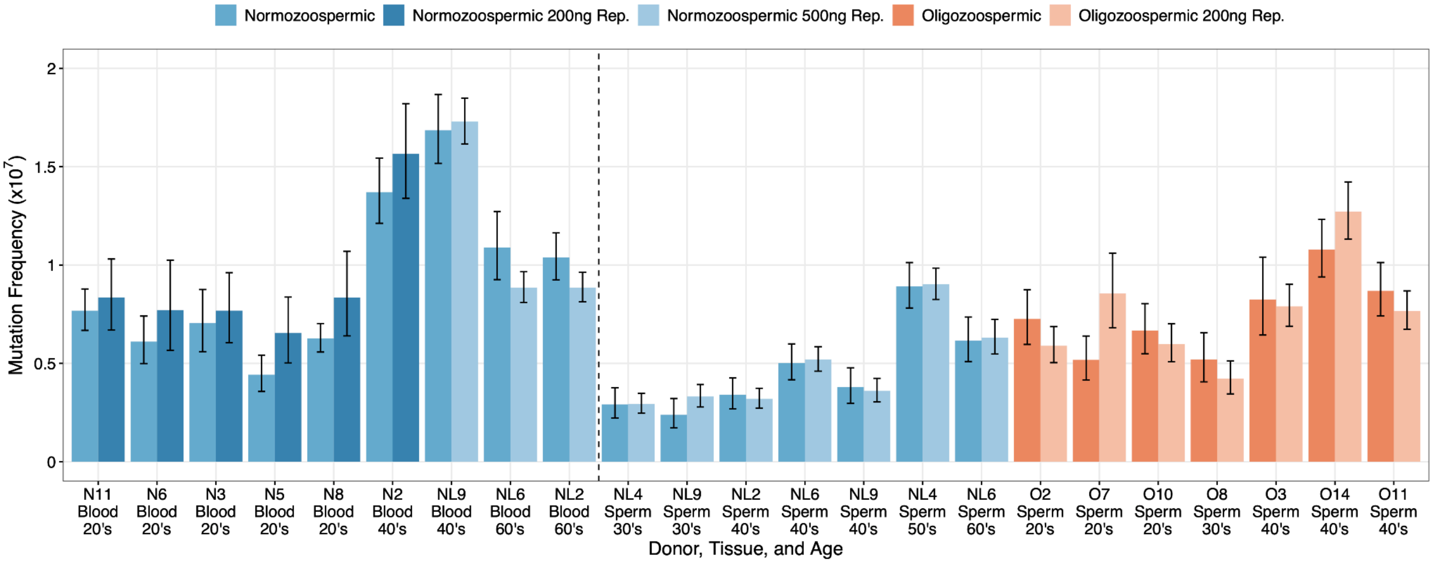


**Supplementary Figure 3**: To demonstrate the consistency of duplex sequencing, we compared mutation rates after extracting and sequencing DNA from variable sperm numbers in fertile controls. This analysis is critical as oligozoospermic men have substantially fewer sperm per ejaculate sample than normozoospermic men. Samples to the left and right of the vertical dotted line are from blood and sperm, respectively. We duplex sequenced technical sperm replicates from n=7 normozoospermic donors using 200ng (blue) and 500ng (lighter blue) of input DNA, equivalent to ~67,000 sperm and ~150,000 sperm used in each replicate library, respectively (**Methods**). We find that mutation rate estimates are consistent across technical replicates with variable DNA input amounts. This result suggests that a sample's germline mutation rate could be precisely determined when sequencing DNA from as few as ~67,000 sperm, empowering the study of sperm mutation in both normozoospermic and oligozoospermic men. Indeed, we further observed consistent mutation rates in technical blood replicates from nine normozoospermic donors (technical replicates sequenced using 200ng and 500ng of DNA) and in seven 200ng technical sperm replicates from oligozoospermic men. S

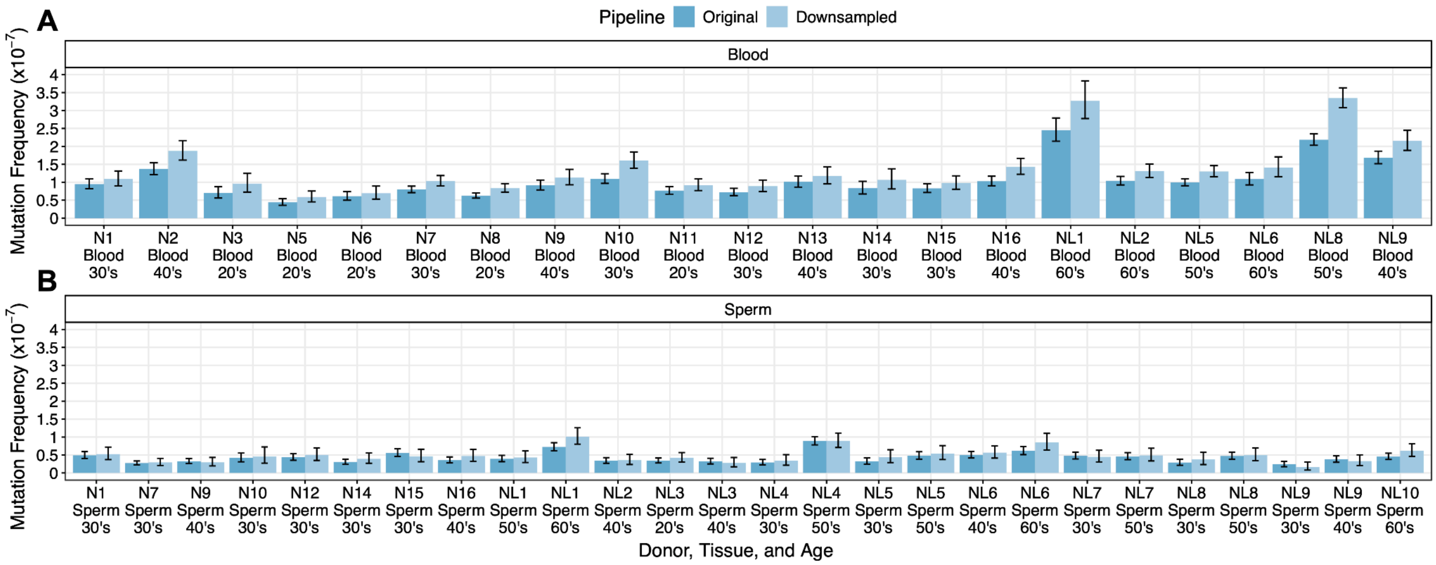


**Supplementary Figure 4**: Mutation frequency estimates in (**A**) blood and (**B**) sperm from normozoospermic men are shown along with 95% confidence intervals when using the original duplex BAM file (darker blue) and the downsampled duplex BAM file (lighter blue). This analysis was done using samples prepared from 200ng of DNA. The downsampling strategy for these sperm and blood samples is outlined in the **Methods** under “Downsampling BAM Files.”

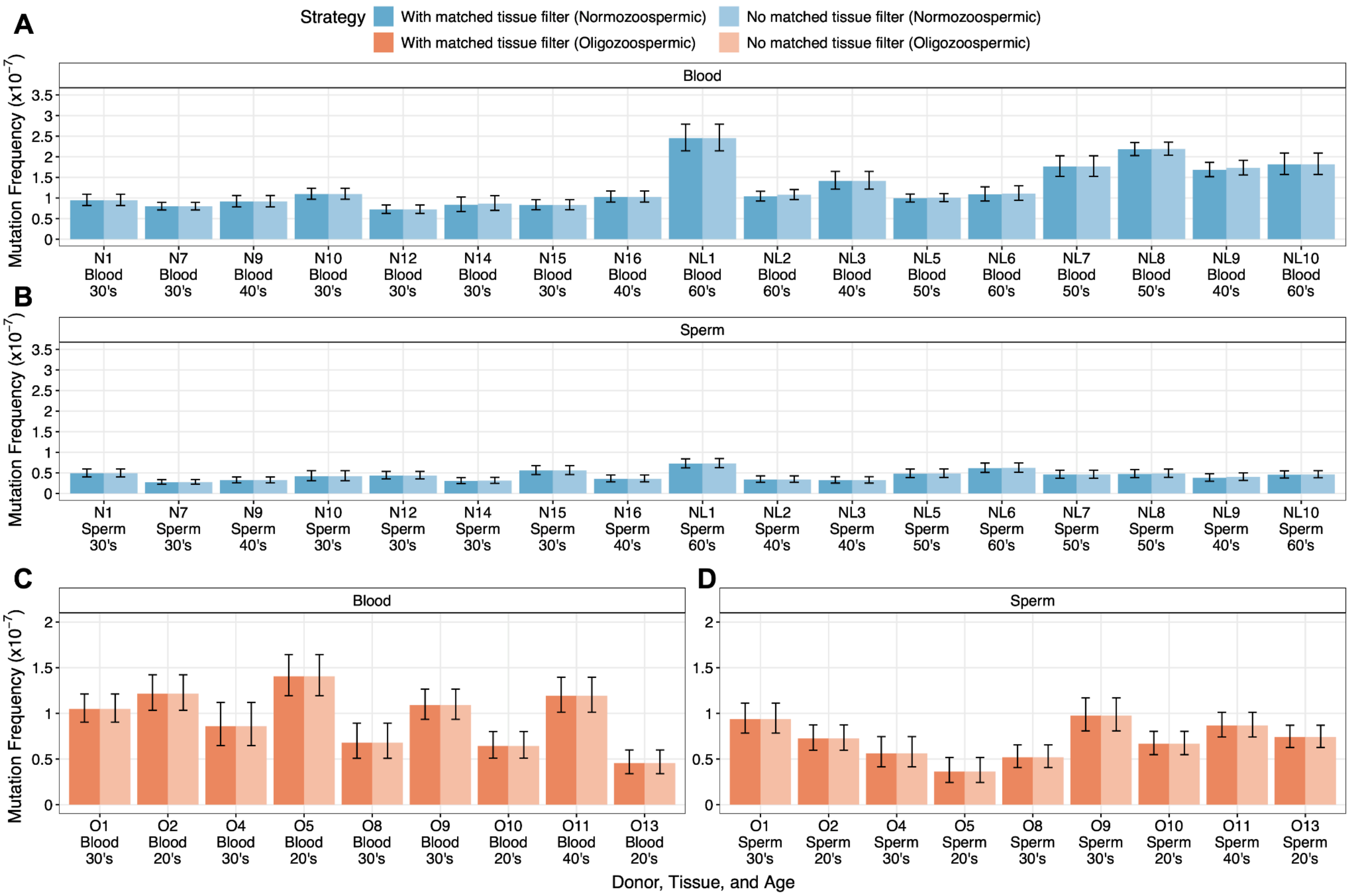


**Supplementary Figure 5**: Mutation frequencies and 95% confidence intervals are shown in normozoospermic (blue) and oligozoospermic (orange) men with successfully prepared 200ng blood (**A** and **C**) and sperm (**B** and **D**) libraries. The darker bar depicts mutation frequency estimates derived from a calculation that employs the within-donor recurrence filter, while the lighter bar depicts measurements when the within-donor recurrence filter was omitted.

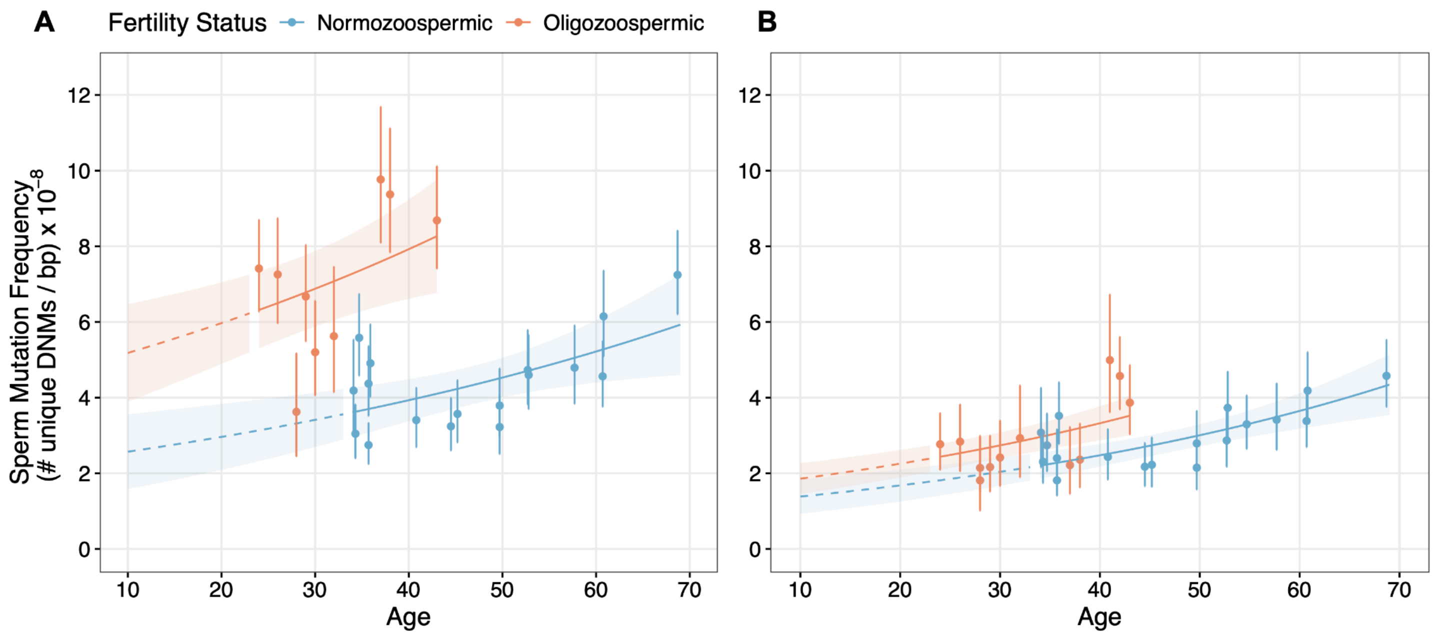


**Supplementary Figure 6**: Mutation frequency estimates and 95% confidence intervals are depicted for sperm samples derived from normozoospermic (blue) and oligozoospermic (orange) donors with (**A**) successfully prepared sperm and blood DNA libraries and (**B**) after excluding all C>T mutations called at non-CpG sites. The blue and orange lines indicate the predicted sperm mutation frequency for a given age based on a negative binomial regression with a log link function. The solid line predicts mutation frequencies at ages within the range of donor ages observed in each cohort. The dashed line predicts values at ages outside of the range of donor ages. The shaded area indicates the 95% confidence interval around the negative binomial regression, calculated by multiplying the standard error by 1.96.

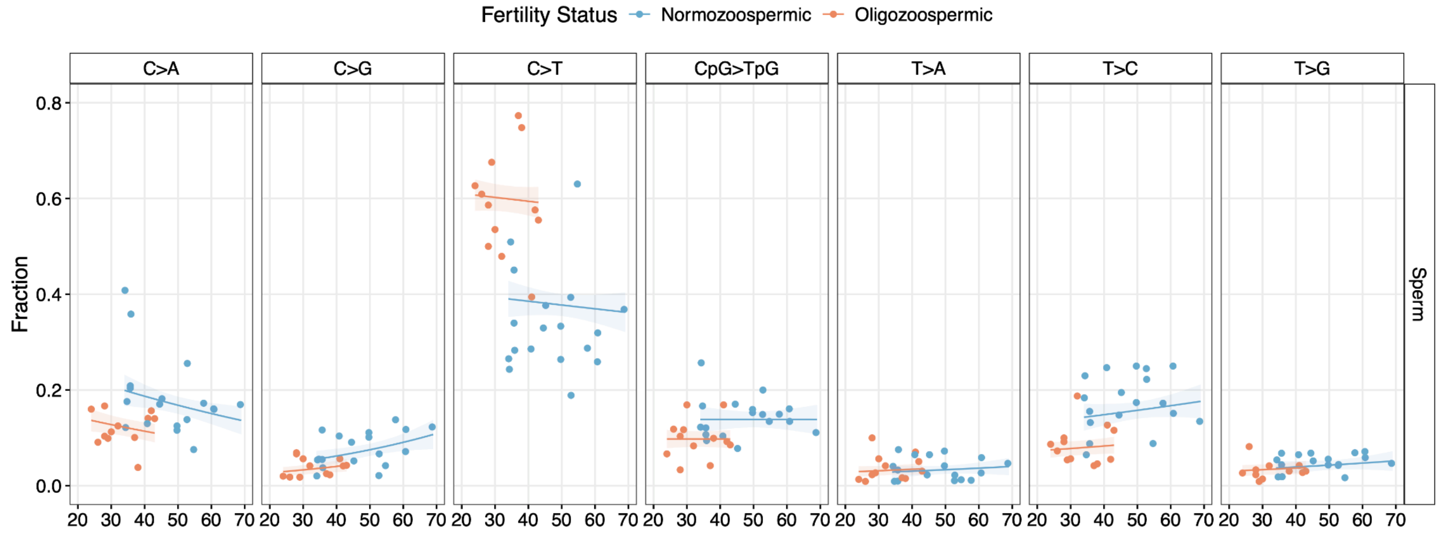


**Supplementary Figure 7**: Binomial regressions with logit links (along with 95% confidence intervals calculated by multiplying the standard error by 1.96) were used to model the fraction of each mutation subtype (C>A, C>G, C>T (non-CpG), CpG>TpG, T>A, T>C, and T>G) across sperm samples in the normozoospermic (blue) and oligozoospermic (orange) cohort. After multiple test corrections (Benjamini-Hochberg procedure), oligozoospermia was significantly associated with an increased C>T (non-CpG) (additive binomial regression $P$ = $5.99 \times 10^{-4}$) and decreased C>A (additive binomial regression $P$ = $7.01 \times 10^{-4}$) and T>C (additive binomial regression $P$ = $7.01 \times 10^{-4}$) fractions.

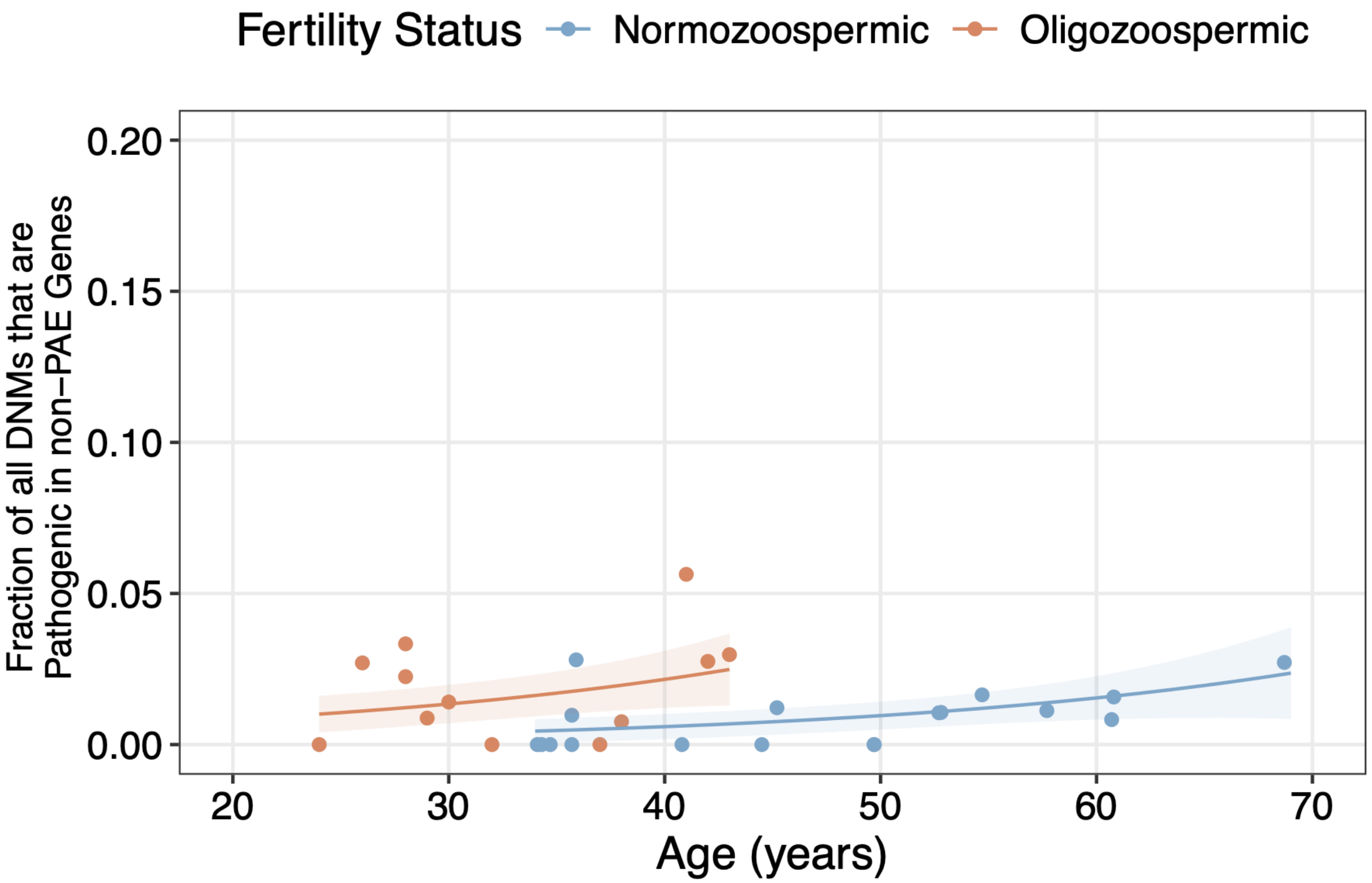


**Supplementary Figure 8**: The fraction of ClinVar pathogenic DNMs (**Methods**) within non-PAE genes measured from 200ng sperm libraries. An additive binomial regression model with a logit link was used to predict the non-PAE pathogenic DNM fraction in normozoospermic (blue) and oligozoospermic (orange) as a function of donor age, sperm motility, and fertility status. The shaded area indicates the 95% confidence interval around the regression model's predicted fraction (1.96 multiplied by the standard error).

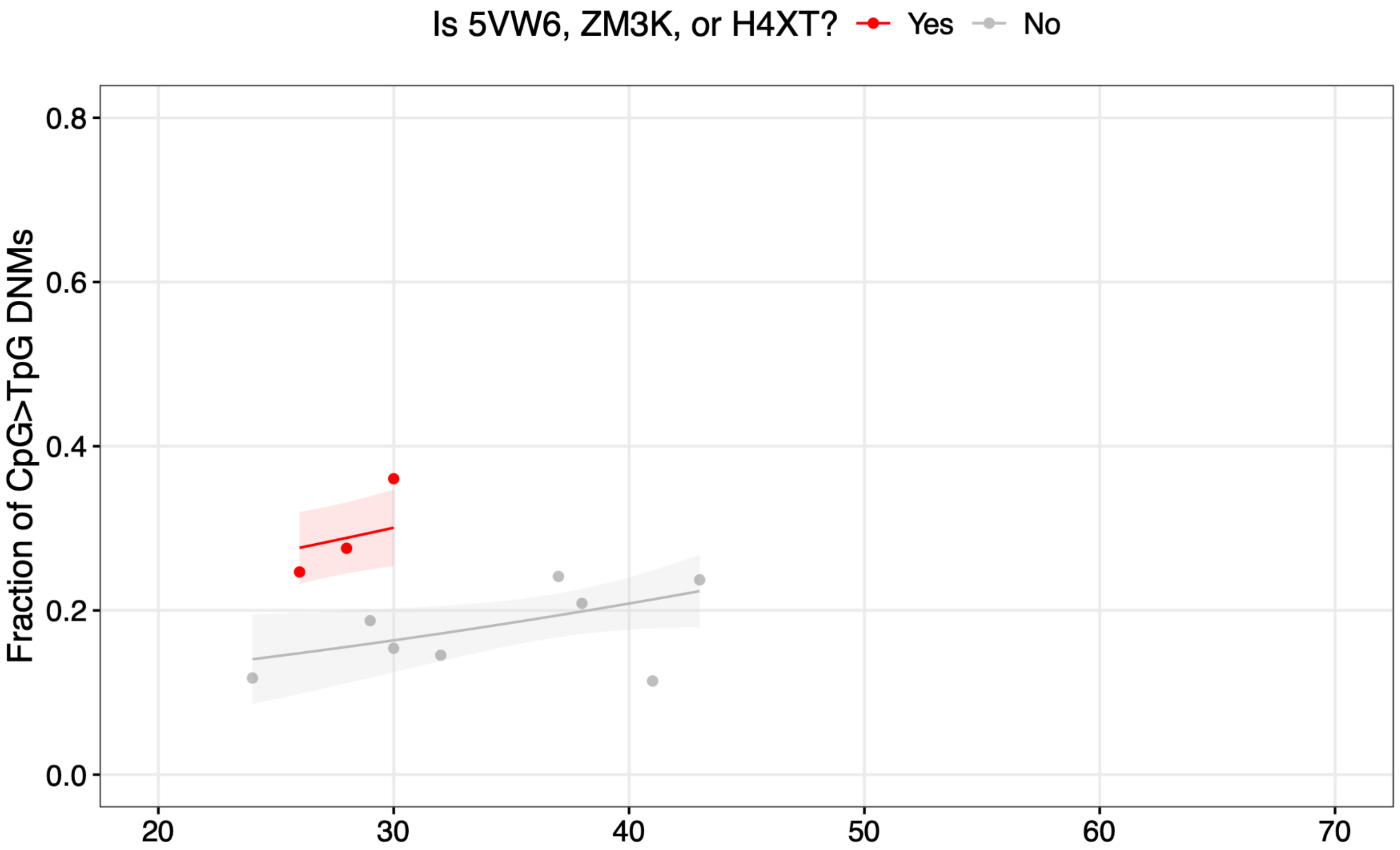


**Supplementary Figure 9**: The fraction of CpG>TpG mutations in each blood sample from oligozoospermic men is plotted along with the predicted fractions measured from an additive binomial regression model with a logit link. The shaded area indicates the 95% confidence interval around the regression model's predicted fraction (1.96 multiplied by the standard error). The CpG>TpG fractions from the three oligozoospermic blood samples that drove our observation for significantly elevated blood mutation frequencies in oligozoospermic men compared to normozoospermic men are shown in red.

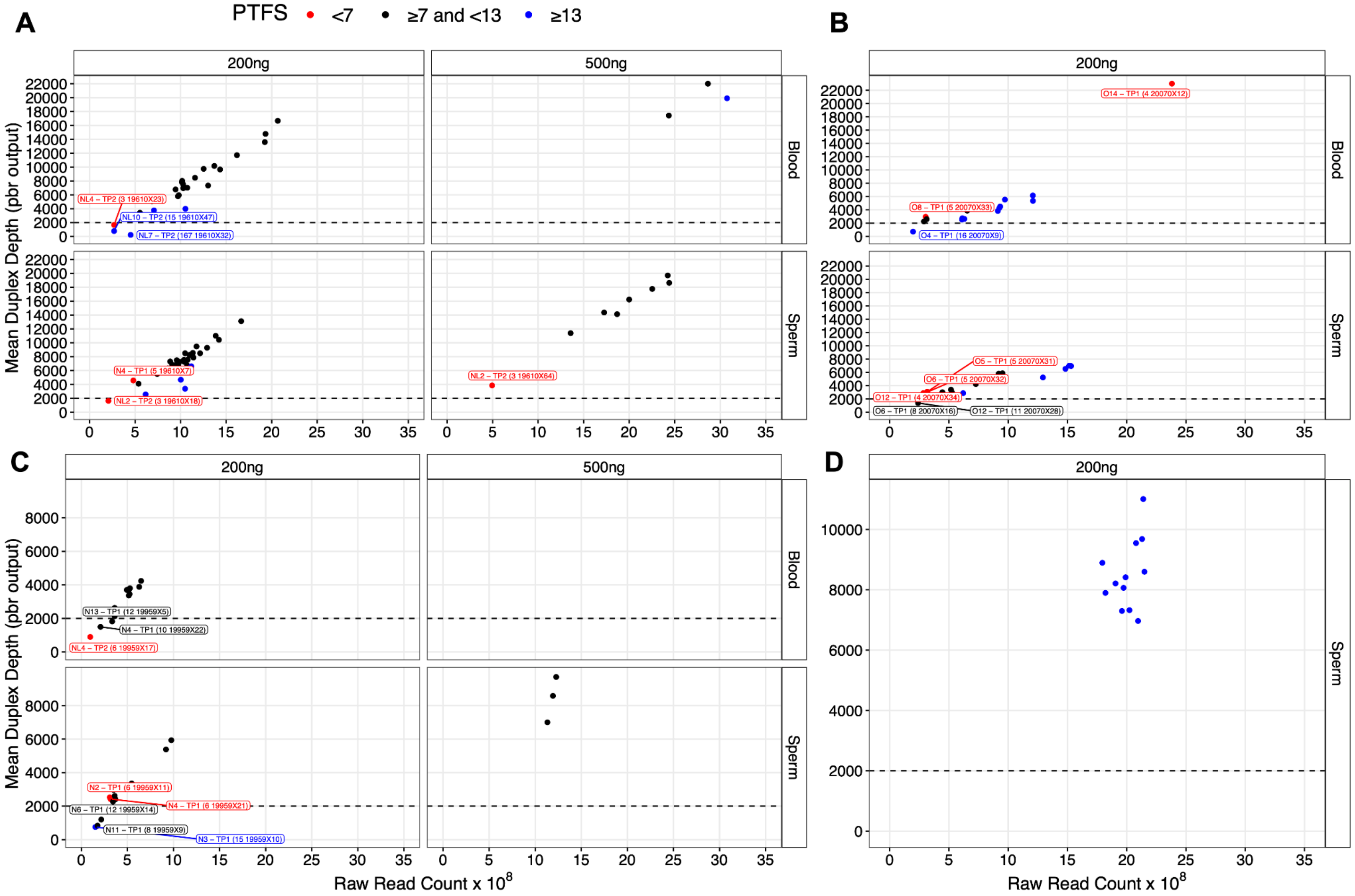


**Supplementary Figure 10**: Mean duplex depth (sum of prefiltered sample depth / 375kb panel) for (**A**) 200ng and 500ng normozoospermic sperm and blood libraries, (**B**) 200ng oligozoospermic libraries, (**C**) 200ng and 500ng normozoospermic sperm and blood libraries that corresponded to samples that were re-prepped and sequenced (e.g., due to library prep failure during the initial experiment), and (**D**) 200ng sperm libraries from normozoospermic and oligozoospermic men whose replicate samples were proceeded in the same batch. The samples depicted in these panels did not exhibit evidence for cross-contamination based on the pair-wise Jaccard index (**Methods**, **Supplementary Figure 11**). The x-axis depicts the total duplex read count for each library. Low-complexity libraries with mean duplex depths ≤2000x are labeled as "Donor ID - time point (PTFS deidentified_sample_ID)." These low-complexity libraries were exclusively used during mutation filtering steps (**Methods**) to flag recurrent mutations across sequencing libraries and were excluded from downstream mutation frequency, spectra, and fraction analyses.

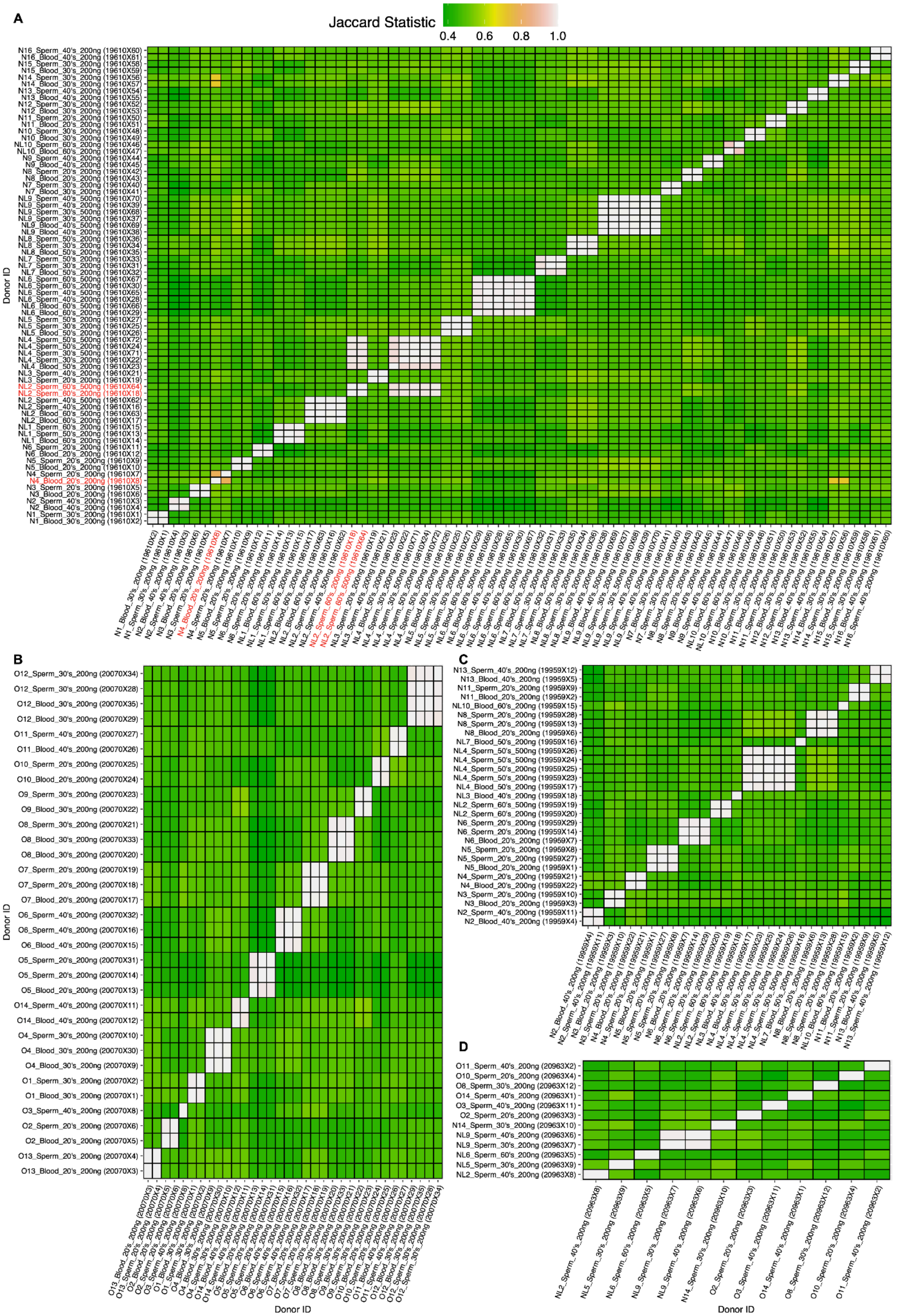


**Supplementary Figure 11**: Using variants with allele frequencies >20%, we calculated the Jaccard statistic (intersection of variants / union of variants) for each pairwise sample combination in the (**A**) normozoospermic cohort, (**B**) oligozoospermic cohort, (**C**) re-prepped and sequenced normozoospermic cohort, and (**D**) replicate normozoospermic and oligozoospermic cohort. Each sample is labeled as “Donor ID_tissue_age_input DNA (sample ID)” such that samples from the same donor should exhibit a Jaccard statistic close to a value of 1 (white cell) due to the widespread sharing of inherited variants. Red-labeled samples exhibited possible cross-contamination in panel A and were subsequently excluded from downstream analyses (outside of performing the recurrence filter).

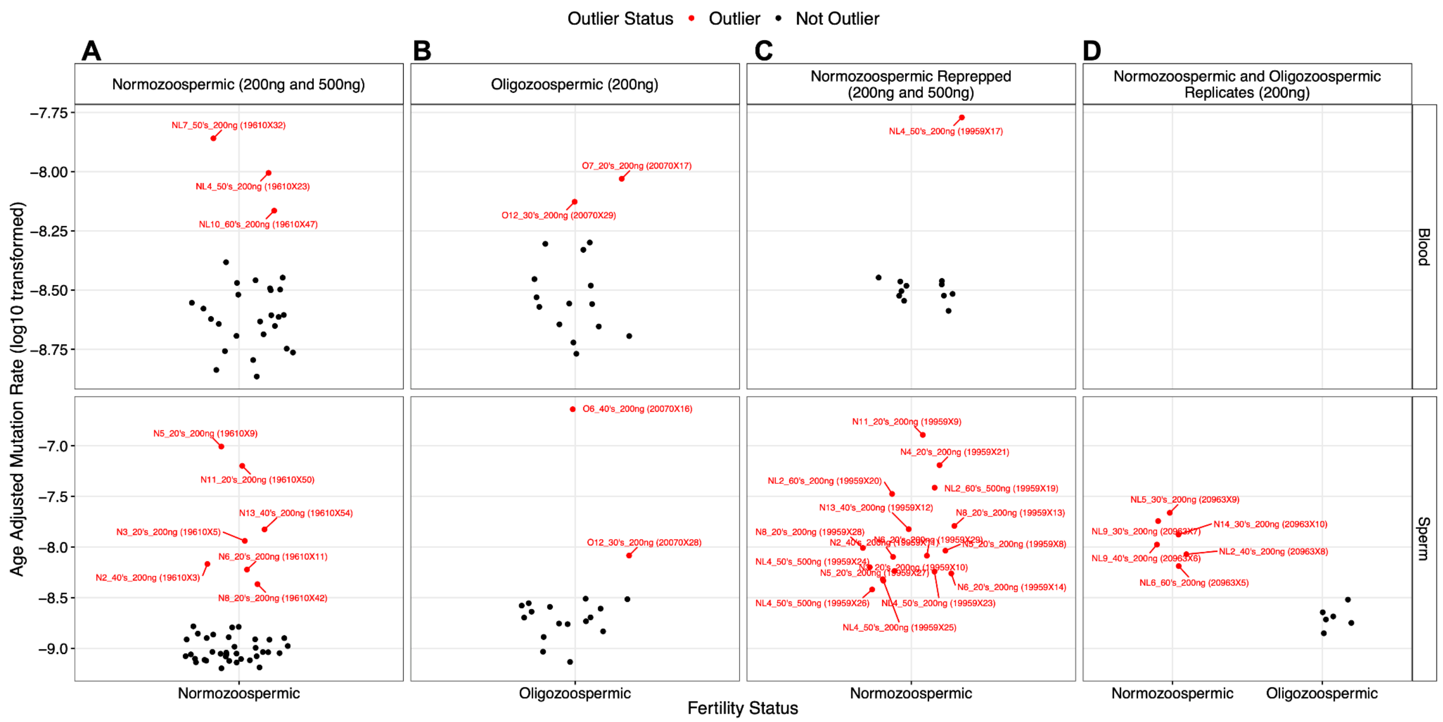


**Supplementary Figure 12**: Age-adjusted mutation frequencies (mutation frequency / donor age) in (**A**) 200ng (non-replicate) and 500ng (replicate) normozoospermic sperm and blood libraries (**B**) 200ng non-replicate oligozoospermic libraries, (**C**) 200ng and 500ng normozoospermic sperm and blood libraries that corresponded to samples that were re-prepped and sequenced (e.g., due to library prep failure during the initial experiment), and (**D**) replicate 200ng sperm libraries from normozoospermic and oligozoospermic men whose samples were proceeded in the same batch. Data points labeled with "Donor ID_age_input DNA (Sample ID)" specify samples with outlier age-adjusted mutation frequency measurements. Outliers in each fertility and tissue group were identified through a median absolute deviation (MAD) test (**Methods**, age-adjusted mutation frequency >3.5 * MAD). Labeled samples were exclusively used to flag recurrent mutations in our variant calling pipeline. Outlier samples in the "Normozoospermic (200ng and 500ng)" group were most significantly enriched for C>A mutations, and outlier samples in the remaining groups were most significantly enriched for C>T mutations (all Chi-square $P < 1\text{x}10^{-200}$).
