## Supplementary figures and images for "Sperm from infertile, oligozoospermic men have elevated mutation rates"

### Supplementary Figure 1 - Sample coverage

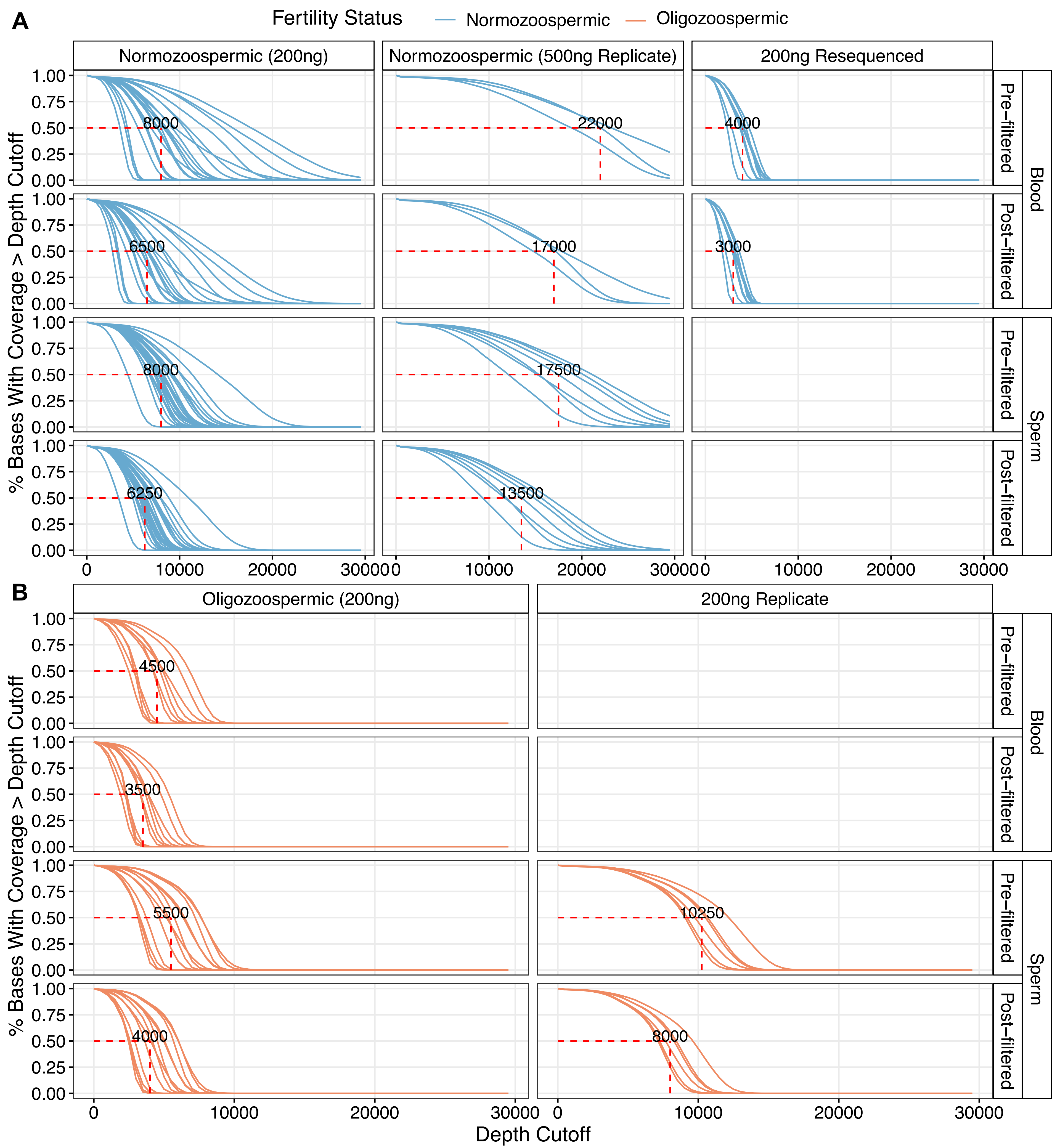

### Supplementary Figure 2 - Nonclonal and clonal DNM fraction

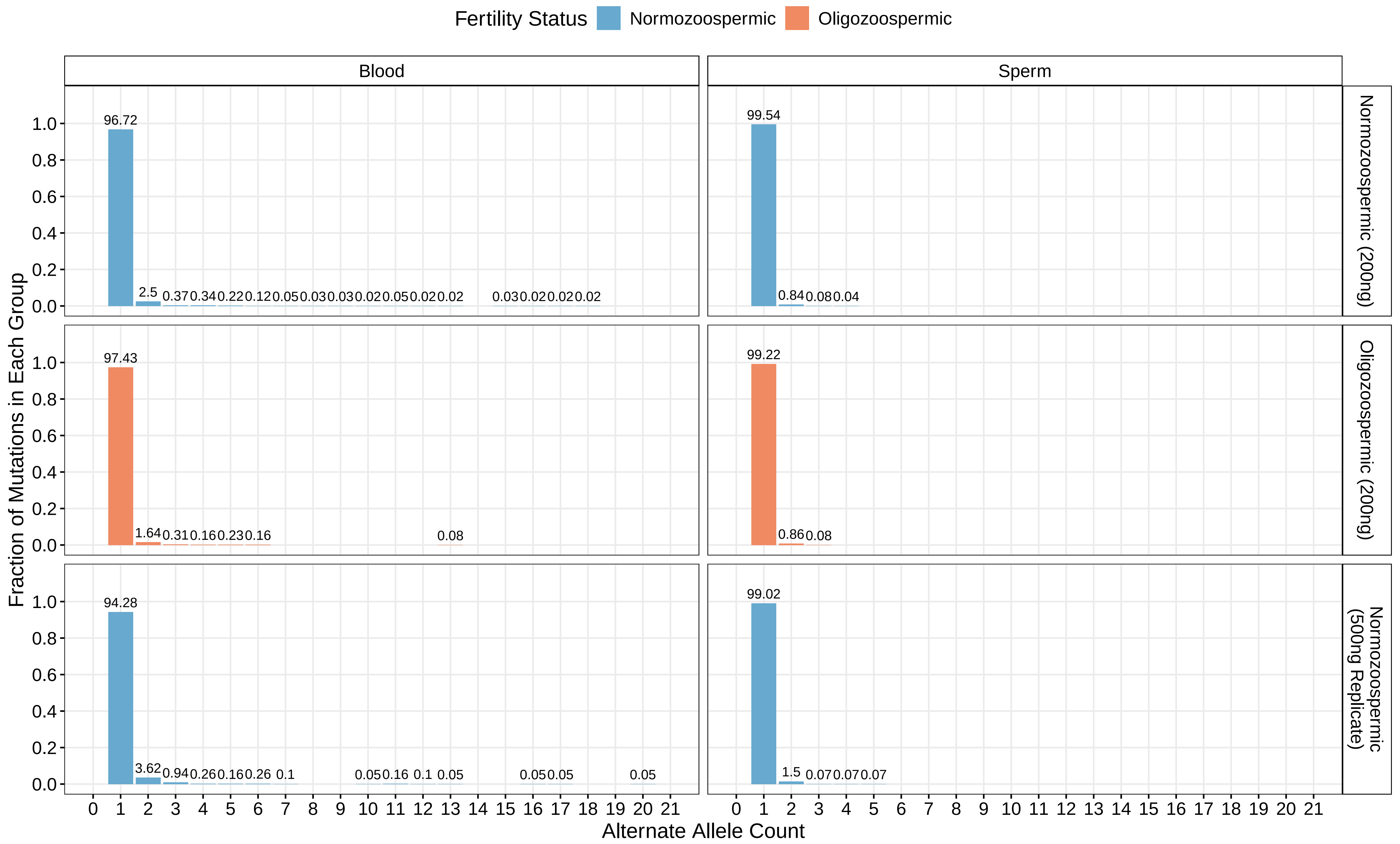

### Supplementary Figure 3 - Mutation frequency in technical replicates

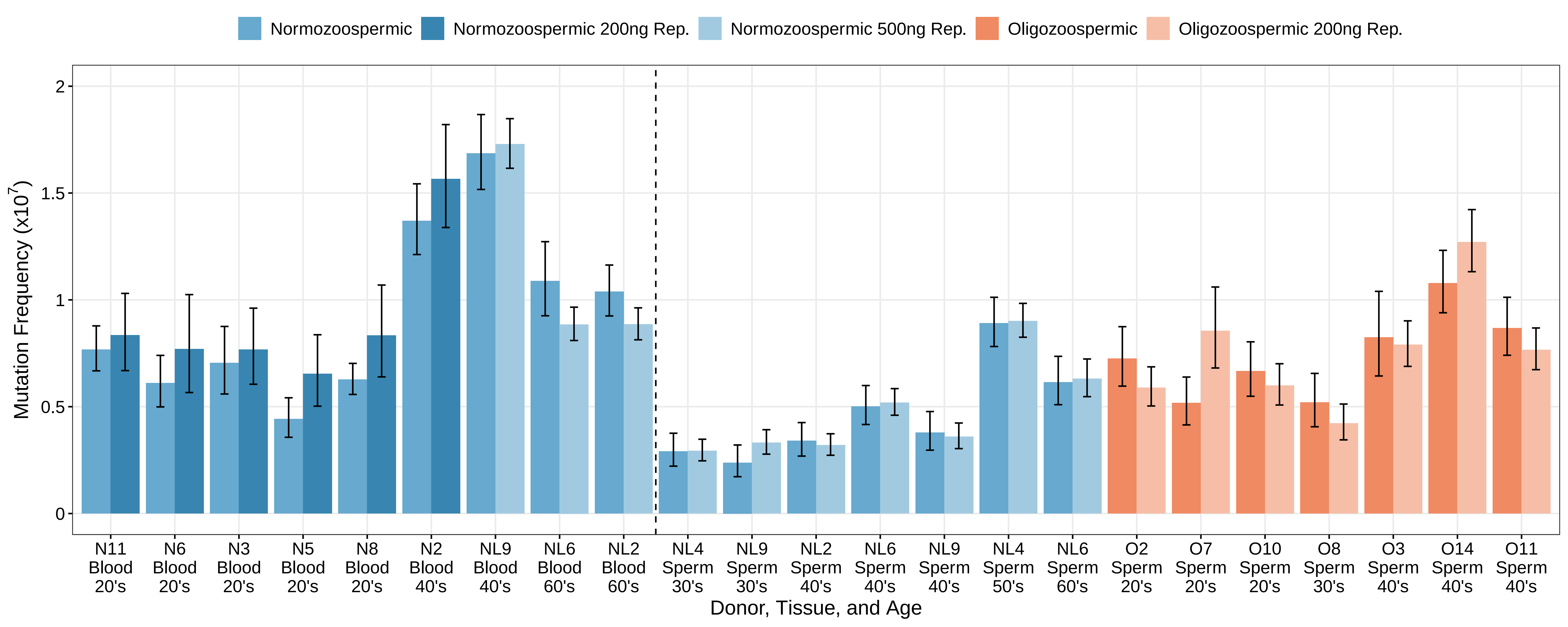

### Supplementary Figure 4 - Downsampled normozoospermic BAM mutation frequencies

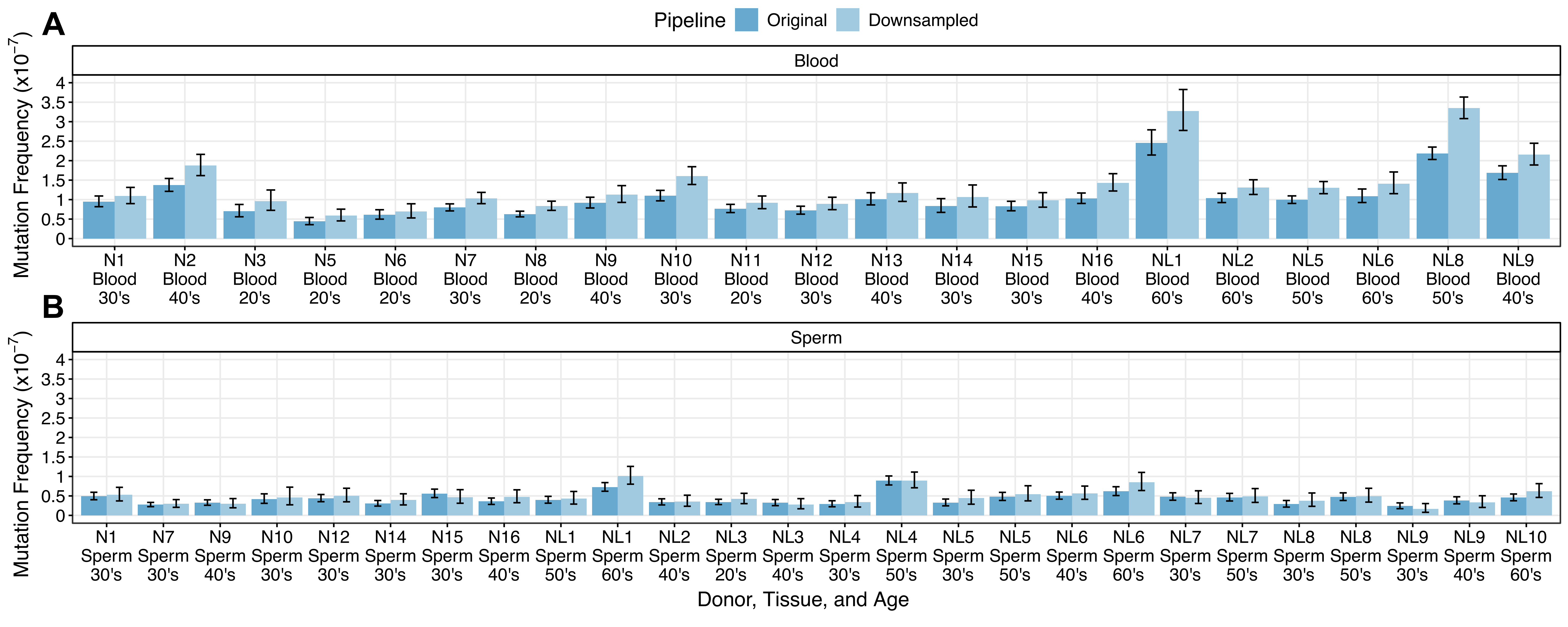

### Supplementary Figure 5 - Matched donor vs recurrence filter mutation frequencies

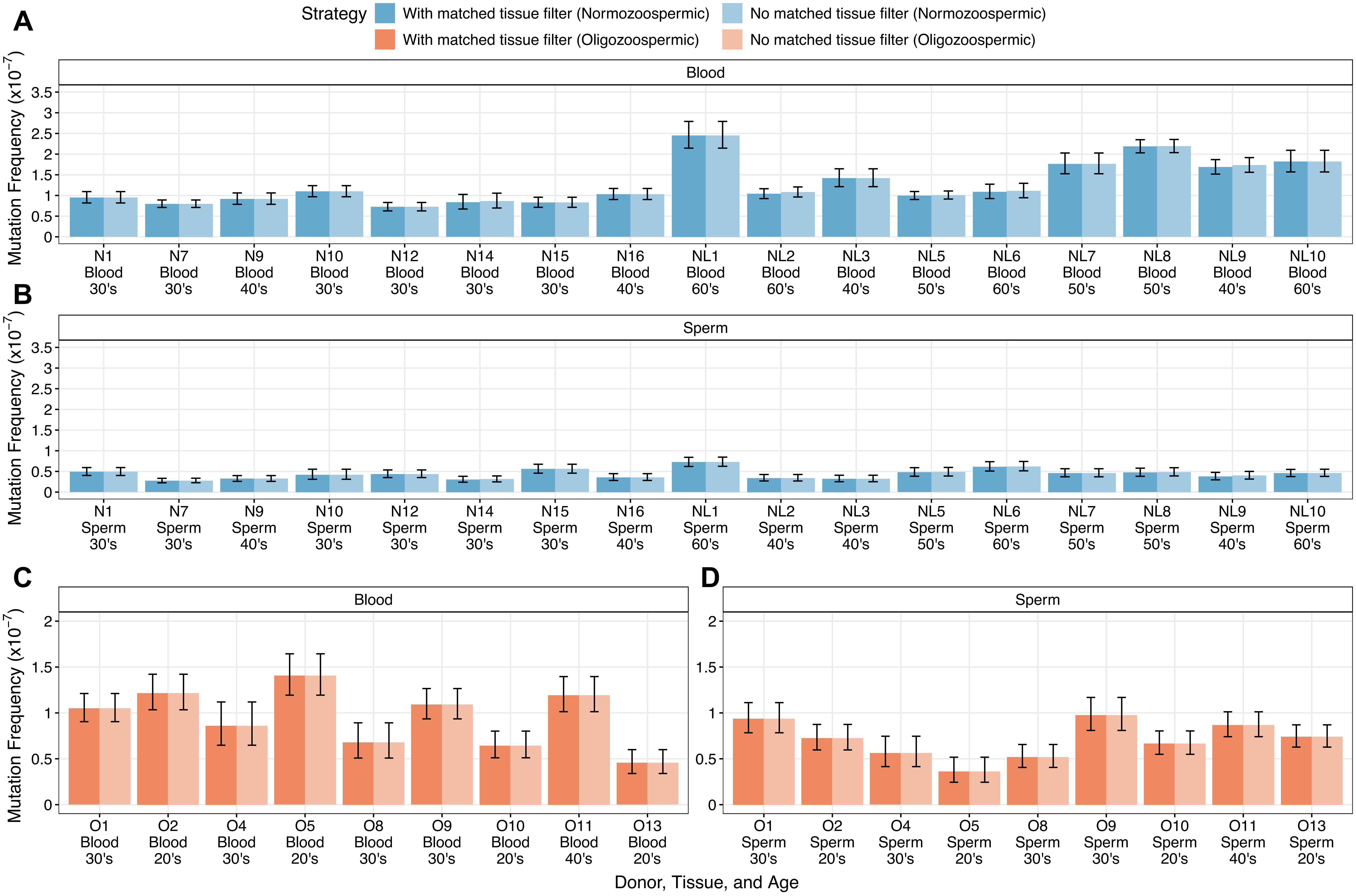

### Supplementary Figure 6 - Matched donor and non-C>T mutation frequencies

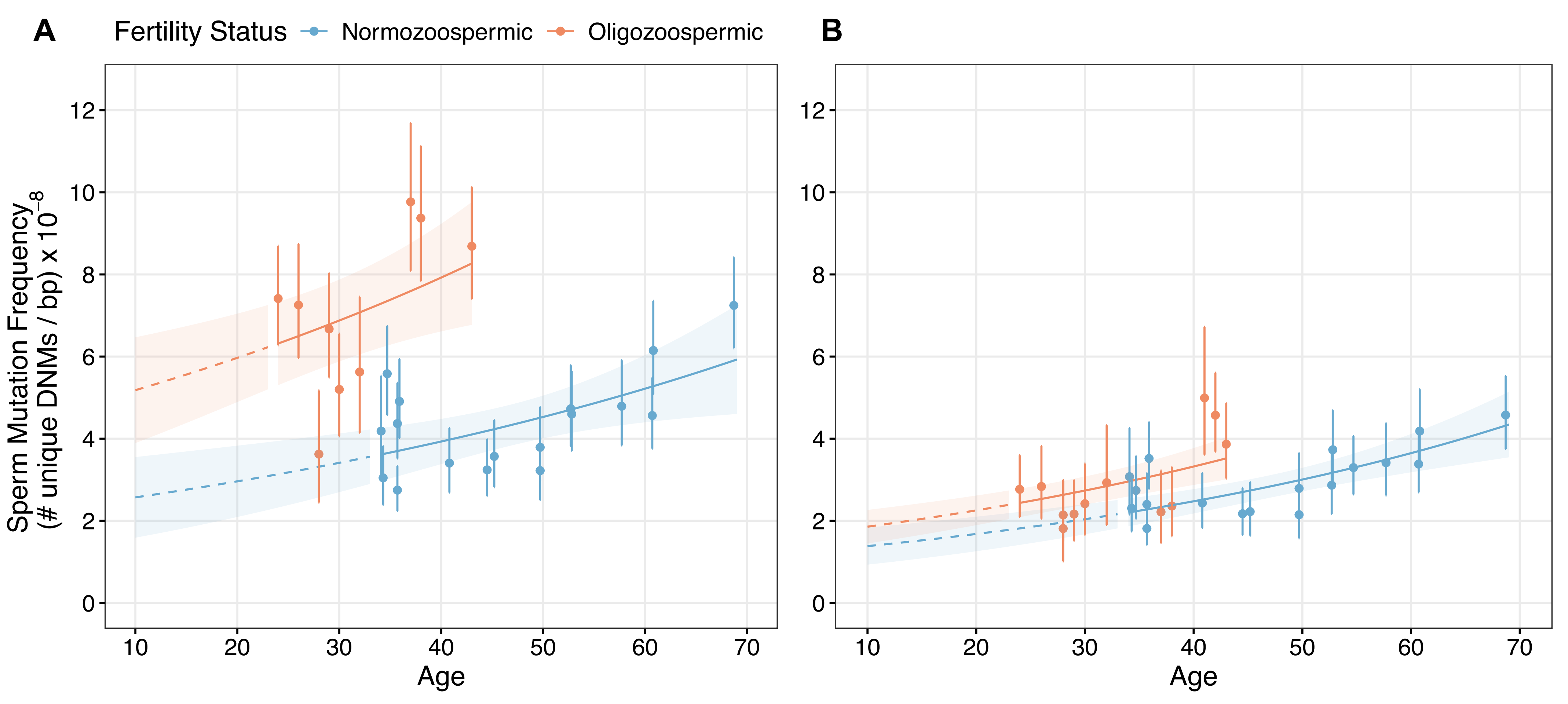

### Supplementary Figure 7 - Sperm mutation spectra

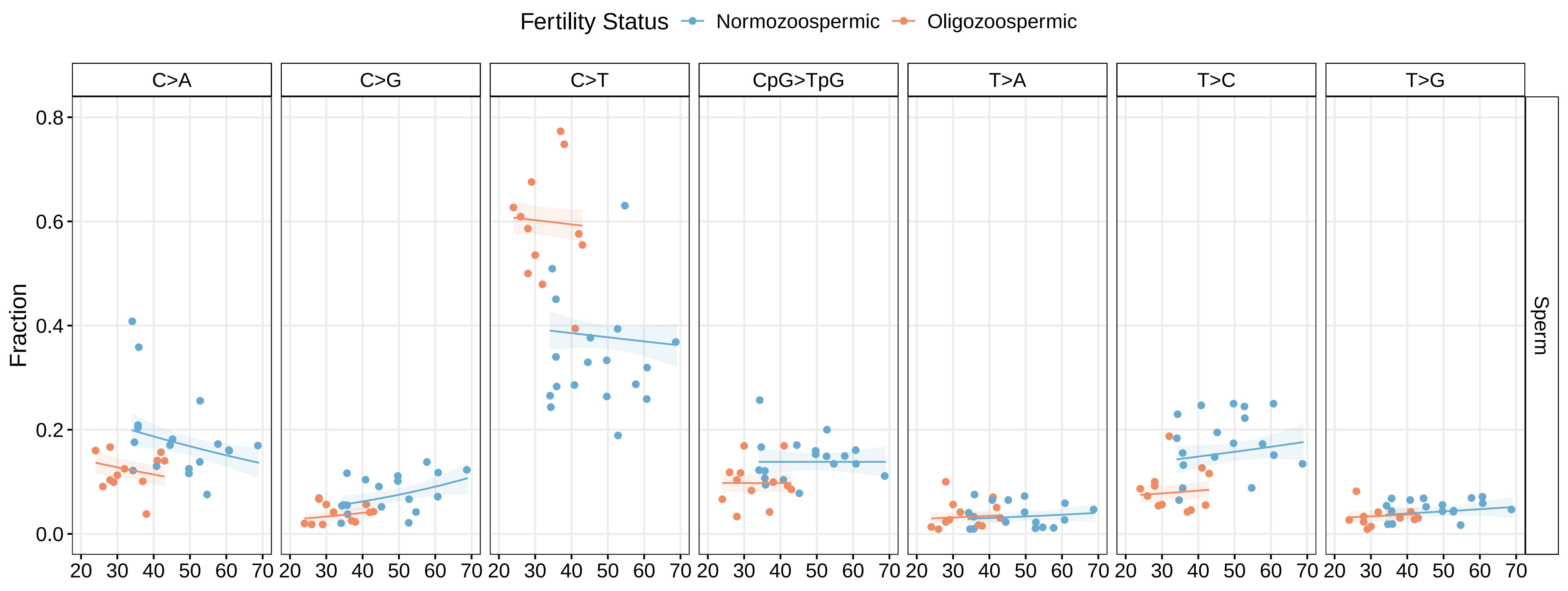

### Supplementary Figure 8 - Fraction pathogenic in non-PAE genes

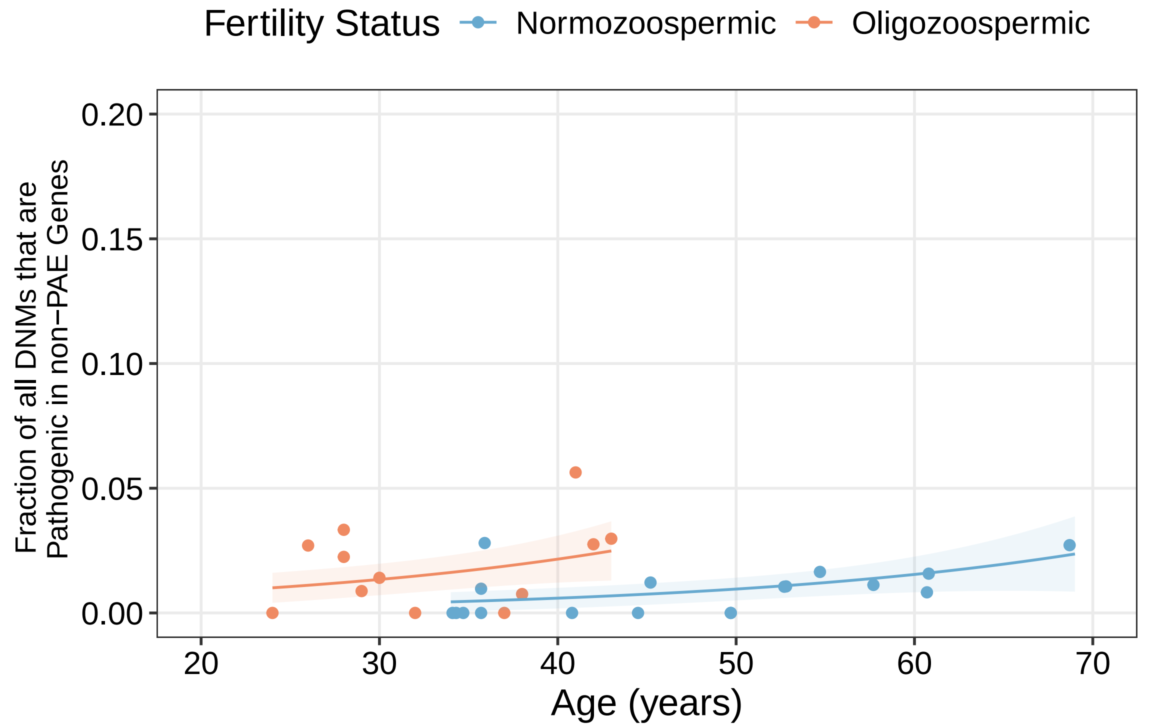

### Supplementary Figure 9 - Blood CpG>TpG spectra

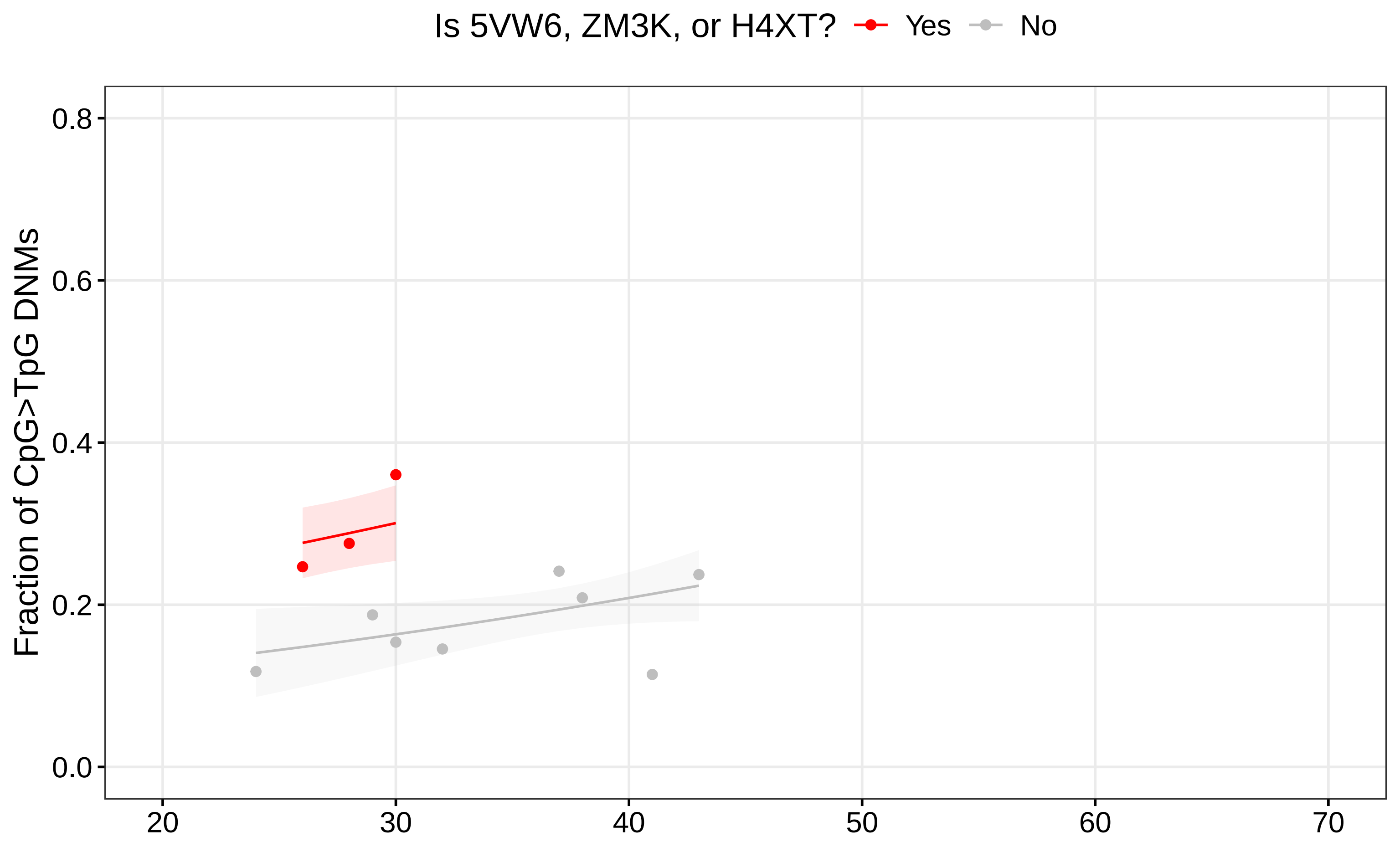

### Supplementary Figure 10 - Library complexity

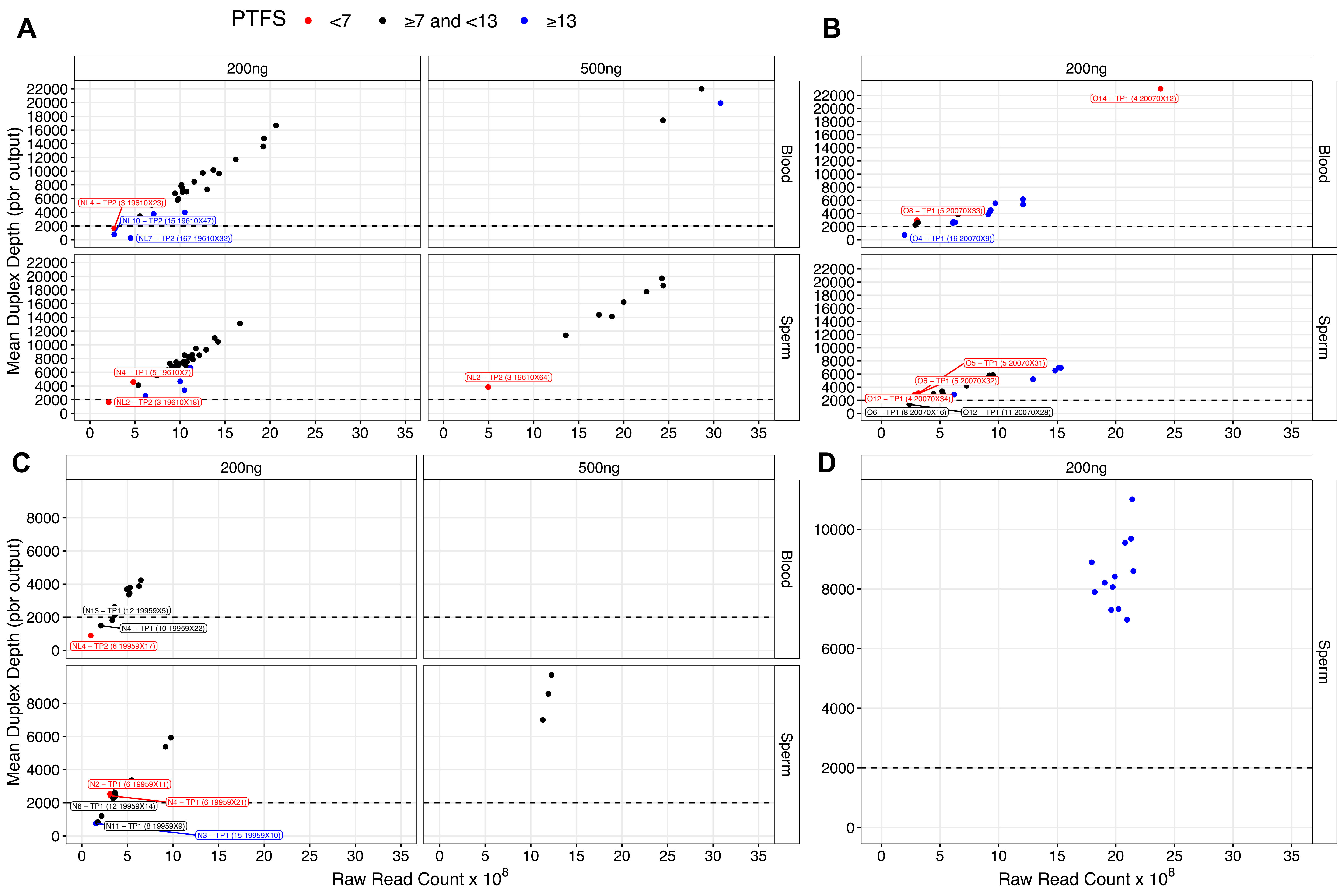

### Supplementary Figure 11 - Jaccard

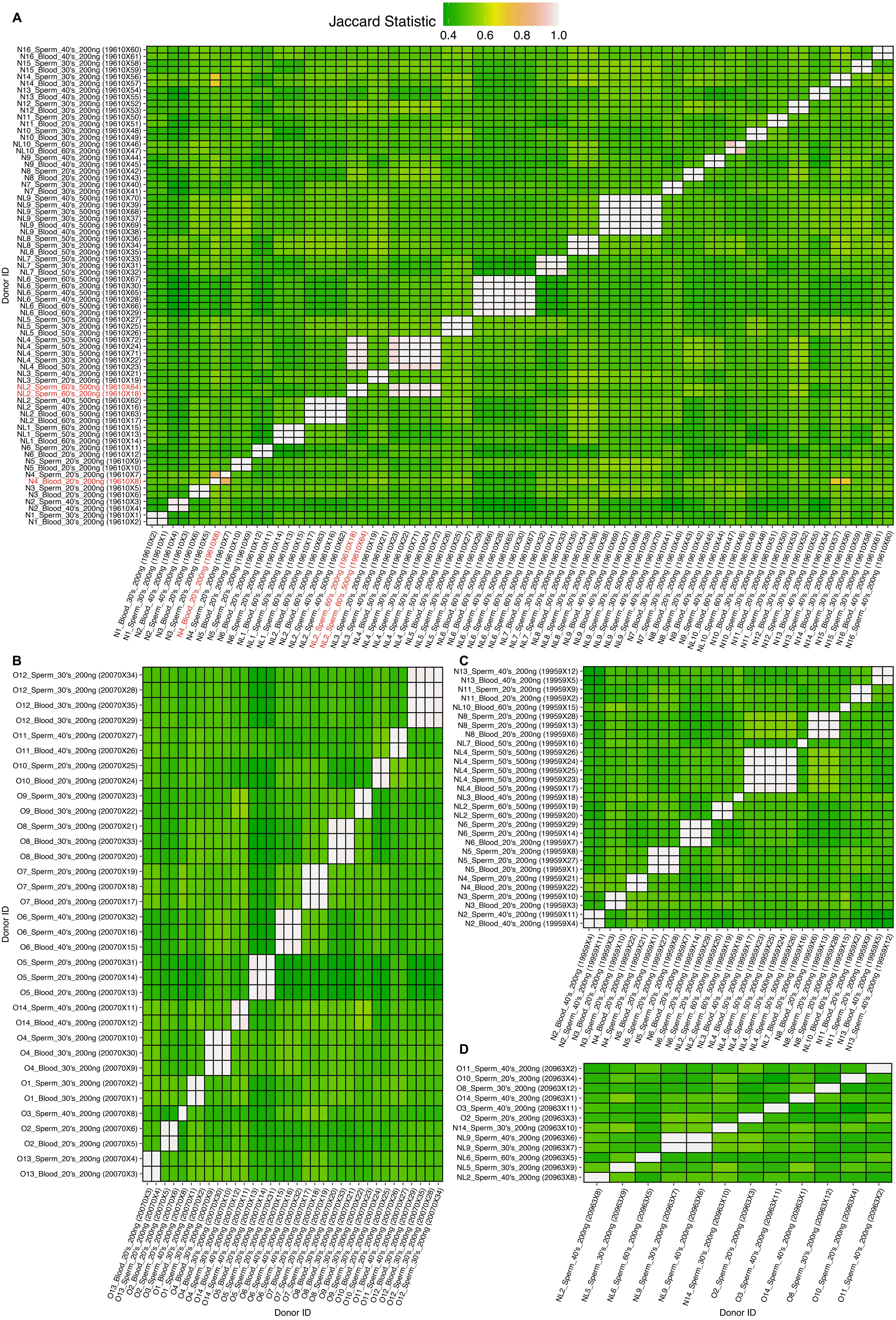

### Supplementary Figure 12 - MAD mutation frequency outliers

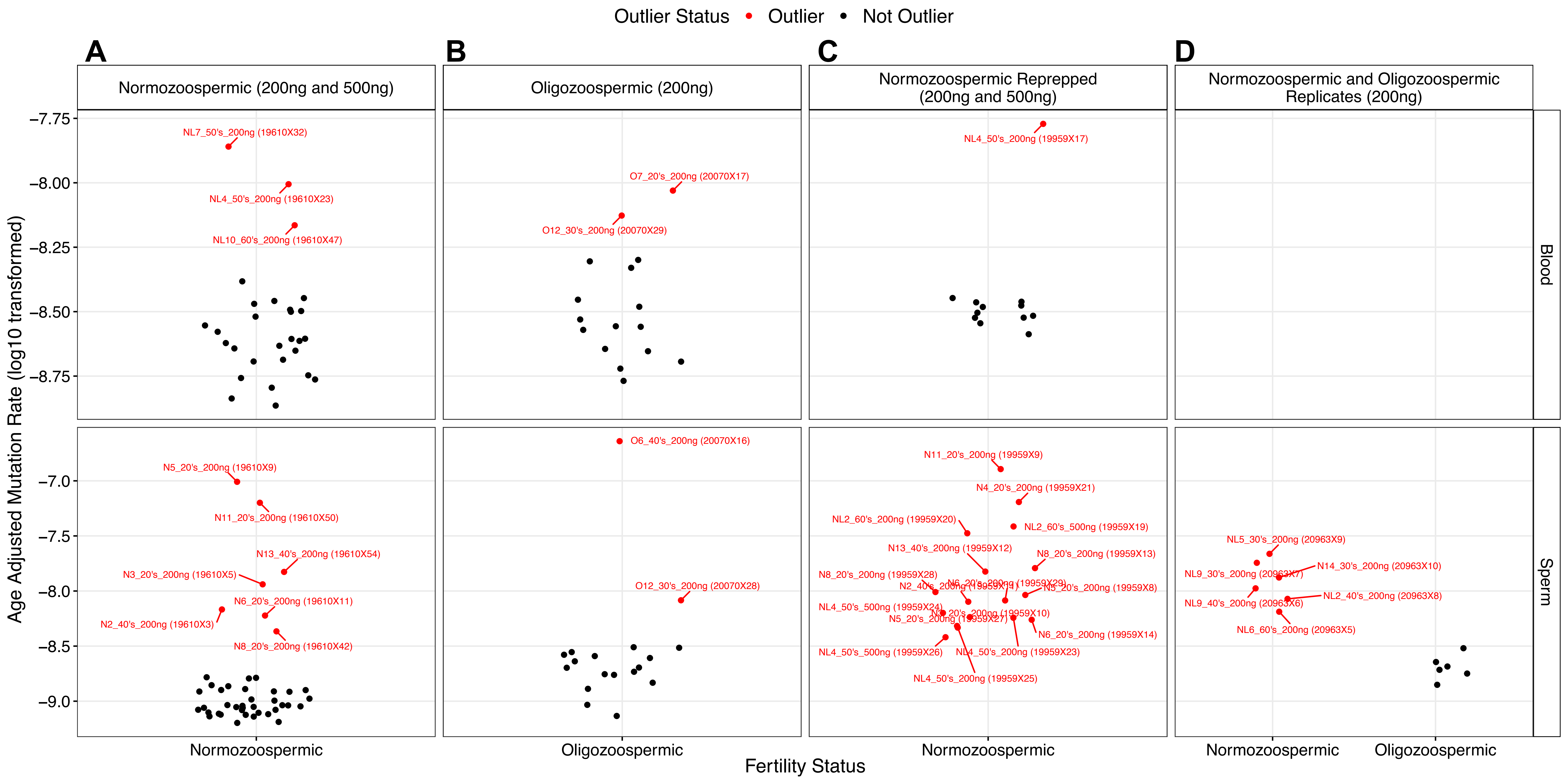
